# Topographic volume-standardization atlas of the human brain

**DOI:** 10.1101/2021.02.26.21251901

**Authors:** Kevin Akeret, Christiaan Hendrik Bas van Niftrik, Martina Sebök, Giovanni Muscas, Thomas Visser, Victor E. Staartjes, Federica Marinoni, Carlo Serra, Luca Regli, Niklaus Krayenbühl, Marco Piccirelli, Jorn Fierstra

## Abstract

**Objective:** Specific anatomical patterns are seen in various diseases affecting the brain. Clinical studies on the topography of pathologies are often limited by the absence of a normalization of the prevalence of pathologies to the relative volume of the affected anatomical structures. A comprehensive reference on the relative volumes of clinically relevant anatomical structures serving for such a normalization, is currently lacking.

**Methods:** The analyses are based on anatomical high-resolution three-dimensional T1-weighted magnetic resonance imaging data of 30 healthy Caucasian volunteers, including 14 females (mean age 37.79 years, SD 13.04) and 16 males (mean age 38.31 years, SD 16.91). Semi-automated anatomical segmentation was used, guided by a neuroanatomical parcellation algorithm differentiating 96 structures. Relative volumes were derived by normalizing parenchymal structures to the total individual encephalic volume and ventricular segments to the total individual ventricular volume.

**Results:** The present investigation provides the absolute and relative volumes of 96 anatomical parcellation units of the human encephalon. A larger absolute volume in males than in females is found for almost all parcellation units. While parenchymal structures display a trend towards decreasing volumes with increasing age, a significant inverse effect is seen with the ventricular system. The variances in volumes as well as the effects of gender and age are given for each structure before and after normalization.

**Conclusion:** The provided atlas constitutes an anatomically detailed and comprehensive analysis of the absolute and relative volumes of the human encephalic structures using a clinically oriented parcellation algorithm. It is intended to serve as a reference for volume-standardization in clinical studies on the topographic prevalence of pathologies.

## Introduction

Many pathologies affecting the brain demonstrate anatomical preferences. This selective vulnerability is referred to as pathoclisis (Vogt and Vogt 1922). Specific anatomical patterns are seen for example in inflammatory conditions such as amyotrophic lateral sclerosis (Bede et al. 2016) or autoimmune encephalitis (Dalmau and Graus 2018). Different brain tumors show distinct topographic anatomical patterns (Akeret et al. 2020). The brain also exhibits an anatomically selective vulnerability to metabolic disease (Patel 1994) or toxic injury (Valk and van der Knaap 1992). Identifying and characterizing such topographic patterns may provide important pathophysiological insights, inform diagnostic criteria and pave the way for anatomically targeted therapeutic approaches. Clinical studies on the topography of pathologies are, however, often limited by the absence of a normalization of the prevalence of pathologies to the relative volume of the affected anatomical structures. Despite numerous imaging-based volumetric studies on brain structures, a comprehensive reference on the relative volumes of clinically relevant anatomical structures serving for such a normalization, is currently lacking.

The present investigation aims to provide an anatomically detailed and comprehensive atlas of the relative volumes of the human encephalic structures. The analyses are based on the anatomical high-resolution three-dimensional T1-weighted MRI data of 30 randomly selected healthy adults (14 females and 16 males) guided by a clinically oriented distinct neuroanatomical parcellation algorithm. It is intended for use in studies analyzing the topographic prevalence of pathologies, where it will allow for volume-standardization. In addition, the supplied parcellation algorithm and dataset provides the basis for a reference database for analyses of the variability of these structures.

## Materials and Methods

### Subjects

Magnetic resonance imaging data of 30 human adult brains were examined. The cohort was composed of 14 females (47%) and 16 males (53%). The subjects were randomly selected from a database of healthy individuals established between 2016 and 2018 (van Niftrik et al. 2018). All subjects were Caucasian, with a mean age of 37.8 (SD 13.0, range 24 to 84) and 38.3 (SD 16.9, range 24 to 70) years, respectively. The mean body height was 166.5 cm (SD 4.0, range 158 to 172) in females and 179.8 (SD 7.7, range 160 to 187) in males. The mean body weight in the female subjects was 55.1 kg (SD 5.1, range 45 to 62) and in the male subjects 79.9 (SD 10.3, range 66 to 108). All females were right-handed, one male was left-handed. Exclusion criteria comprised an age below 18 years as well as the presence or a previous history of a medical or neurological disease, neurological symptoms, traumatic brain injury or psychiatric diagnosis. Prior to inclusion in the study, all participants gave their written, informed consent. The study was approved by the local ethics committee (KEK ZH 2012-0427).

### Data acquisition

All subjects underwent the same imaging protocol. A 3-tesla Skyra VD13 (Siemens Healthcare, Erlangen, Germany) with a 32-channel receive coil was used for MRI data acquisition. An anatomical three-dimensional high-resolution T1-weighted Magnetization Prepared Rapid Acquisition Gradient Echo (MPRAGE) image was acquired with the following specification (van Niftrik et al. 2018): ACPC line plus 20° (on a sagittal image), voxel size 0.8 × 0.8 ×1.0 mm^3^, field of view 230 × 230 × 176 mm^3^, scan matrix of 288 × 288 × 176, TR/TE/TI 2200/5.14/900 ms, flip angle 8°.

### Segmentation and volumetric analysis

For volumetric analysis, a hybrid approach with combined manual and semi-automated anatomical segmentation was used. *Supplemental Table 1* shows the chosen method per structure. Semi-automated segmentation was performed for selected structures, for which the high reliability of such an approach has been proven. The remaining structures were segmented manually. Semi-automated volumetric segmentation was done using Freesurfer 5.3.0 (http://surfer.nmr.mgh.harvard.edu/fswiki/FreeSurferWiki)(Fischl et al. 2002, 2004). The anatomical T1-weighted MPRAGE images were automatically segmented into different anatomical subregions according to the Destrieux atlas (file name: aparcþaseg2009s.mgz)(Destrieux et al. 2010; Hendrik Bas van Niftrik et al. 2020). Anatomical regions of interest (ROI) were derived from the subjects’ specific tissue parcellation. The total volume of the cerebral gyral segments was determined by the sum of the respective cortical and white matter areas. The corresponding gyral segments served in their sum to calculate the lobar volumes. For manual segmentation, the medical DICOM imaging software OsiriX MD™ 11.0 was used. Following the structural boundaries defined by the neuroanatomical parcellation algorithm (see below), the corresponding ROI were identified using the appropriate planes in the MPRAGE images (*Supplemental Figure 1*).

### Neuroanatomical parcellation

The parcellation of the encephalon was performed according to the anatomical algorithm described in *Supplemental Table 1* and illustrated in *Figures 1–5*. The anatomical nomenclature follows the *Terminologia Anatomica* and *Terminologia Neuroanatomica* (Ten Donkelaar et al. 2017) with the distinction of a central and a limbic cerebral lobe (Yasargil 1994; Akeret et al. 2019, 2020). Structural delineation was based on neuroanatomical landmarks described in multiple atlases (Lorente De Nó 1934; Carpenter 1972; Salamon et al. 1976; Mai et al. 2008; Nieuwenhuys et al. 2008; Duvernoy 2012; Mai and Paxinos 2012; Duvernoy et al. 2013).

**Figure 1.**
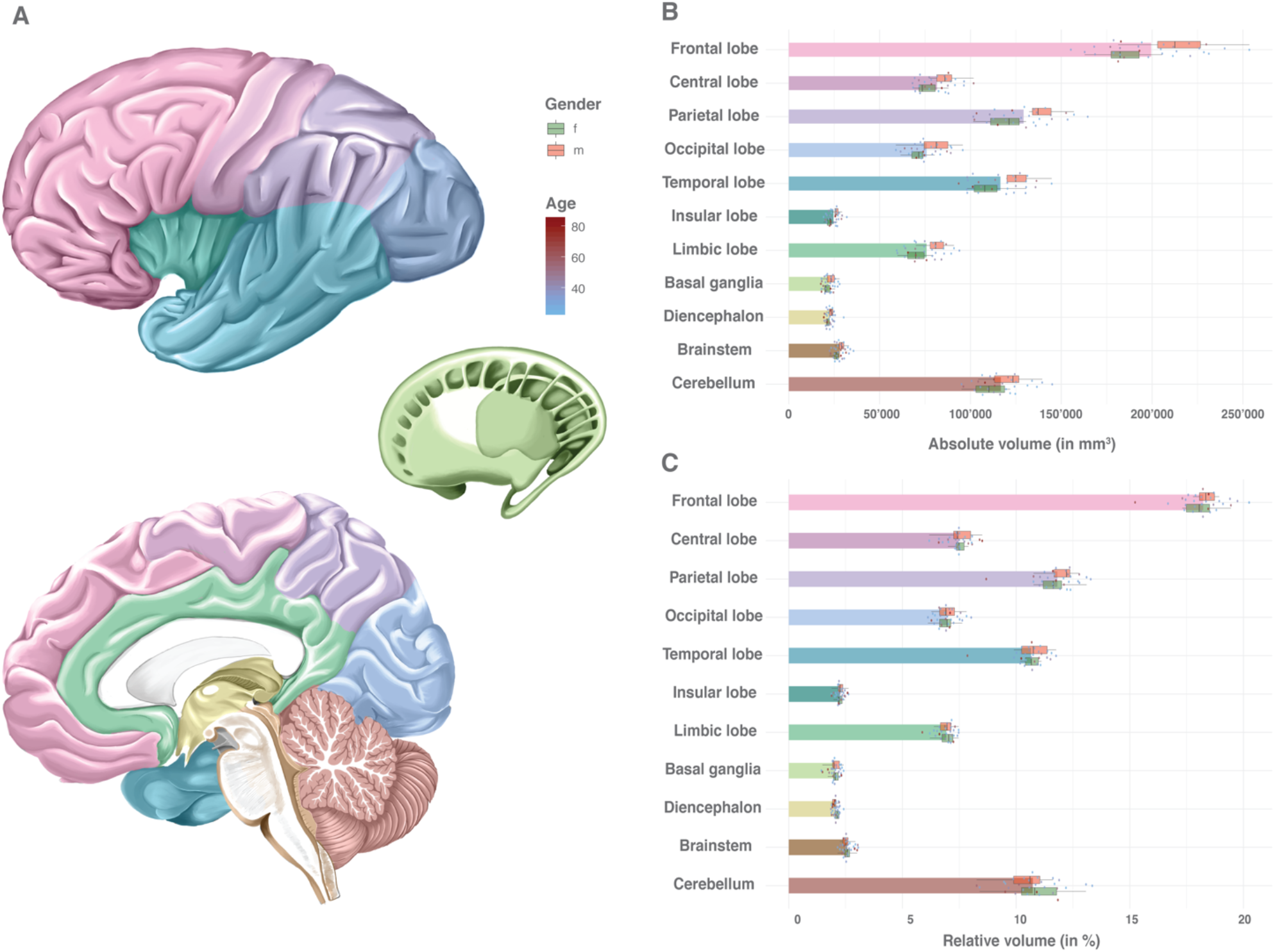
Volumes of general anatomical structures: Absolute and relative volumes of the cerebral lobes, the basal ganglia, the diencephalon as well as the brainstem and cerebellum. **A:** Color scheme of the anatomy, gender and age. **B:** Absolute volumes in mm^3^. Mean volumes are given as bars. Gender stratification is provided with boxplots. The individual measurements are shown as dots with continuous color-coding indicating age. **C:** Relative volumes normalized to the total individual encephalic volume (without ventricles). The graphic elements follow B. *Abbreviations*: f = female; m = male.

### Statistical analysis

Categorical data are given as absolute numbers and percentages (%), continuous data as mean and standard deviation (SD). The SD of absolute and relative volumes are further reported in proportion to the mean as relative standard deviation (RSD). Corresponding to the coefficient of variance, the RSD permits a comparison of the variance between absolute and relative volumes. In view of varying needs concerning anatomical details, the reported volumes are divided into a topographic overview, cerebral gyral segments, central prosencephalic structures, brainstem and cerebellum, and the ventricular system. The relative volumes are given in relation to the total individual encephalic volume (parenchymal, i.e. without ventricular system) for each parcellation unit, and in relation to anatomically appropriate reference volumes in selected cases (e.g. mesencephalon relative to the total individual brainstem volume; subunits of the cerebellum relative to the total individual cerebellum; divisions of the ventricular system relative to the total individual ventricular volume). Gender-specific differences in absolute and relative volumes were analyzed using a two-sample t-test. Pearson correlation was used to assess the association between volumes and age. No significance level was defined; instead, the results were qualitatively interpreted based on the level of evidence: p < 0.001: very strong evidence; p < 0.01: strong evidence; p < 0.05 evidence; p < 0.1 weak evidence; p > 0.1: no evidence(Bland 2015). All statistical analysis was performed using R 4.0.0 (R Core Team 2020).

### Data availability

The authors confirm that the data supporting the findings of this study are available within the article and its supplemental materials. In addition, the anonymized complete dataset is provided as a basis for further investigations in the *Supplemental Raw Data*.

## Results

The mean total encephalic volume without ventricles was 1 093 437 mm^3^ (SD 111 353 mm^3^, RSD 10.2%, range 921 292 mm^3^ to 1 274 242 mm^3^). A significant difference was seen between genders (*females*: mean 1 024 922 mm^3^ (SD 89 785, RSD 8.8%); *males*: mean 1 153 388 mm^3^ (SD 93 653, RSD 8.1%); *p = 0*.*001*). The relation of total encephalic volume without ventricles to subject height and body weight are shown color-coded by gender in *Supplemental Figure 2A*. Despite a trend towards lower volumes with increasing age, no evidence for a correlation was found (p = 0.41, *Supplemental Figure 2B*).

### Topographic overview

The absolute and relative volumes of the different cerebral lobes, the basal ganglia, the diencephalon, the brainstem and the cerebellum are given in *Supplemental Table 2* and *Figure 1*. Among the cerebral lobes, the frontal lobe was the most voluminous (18.2%, SD 0.96, RSD 5.3%). With a relative volume of 2.3% (SD 0.18, RSD 8.2%), the insular lobe was the smallest one. The cerebellum accounted for 10.7% (SD 1.17, RSD 10.9%) of the total encephalic volume. While all anatomical structures had a higher volume in males than in females, no differences between genders remained after normalization to the total individual encephalic volume. The exception to this was the diencephalon, for which some evidence of a gender difference in relative volumes was observed (p = 0.045). The RSD in absolute volume was comparable for all structures. The normalization to the individual total encephalic volume resulted in a distinct reduction in the relative variance. Female and male subjects showed a comparable RSD in both absolute and relative volumes. A general trend for decreasing volumes with increasing age was observed, but without evidence for a significant correlation (*Supplemental Figure 3*).

### Cerebral gyral segments

*Supplemental Table 3* and *Figure 2* show the absolute and relative volumes of the cerebral gyral segments. For detailed data on separate cortical and white matter volumes for each gyral segment, please refer to the *Supplemental Raw Data*. The most voluminous gyri were the superior frontal (6.3%, SD 0.39, RSD 6.2%), the middle frontal (5.3%, SD 0.32, RSD 6.0%) and the precentral gyrus (3.6%, SD 0.30, RSD 8.4%), followed by the parietal gyral segments (angular gyrus (3.1%, SD 0.30, RSD 9.5%), superior parietal lobule (3.05%, SD 0.30, RSD 9.8%), precuneus (2.8%, SD 0.22, RSD 7.8%) and supramarginal gyrus (2.8%, SD 0.30, RSD 10.6%)). The subcallosal area (0.22%, SD 0.0047, RSD 21.7%) showed the smallest volume, with the planum temporale (0.28%, SD 0.057, RSD 20.3%), the anterior orbital (0.31%, SD 0.037, RSD 12.2%) and the rostral gyrus (0.33%, SD 0.025, RSD 7.6%) following in ascending order. All gyral segments demonstrated larger volumes in males than in females. This gender-effect was compensated for by normalization to the total individual encephalic volume. The RSD of absolute volumes demonstrated an upward trend with decreasing absolute volume of the respective structure. The largest relative variance was observed in the gyrus rectus (45.9%), the subcallosal area (27.7%), the planum temporale (25.0%), planum polare (18.7%), long insular (20.6%) and the orbital gyri (18.0-18.2%). By normalizing to the total individual encephalic volume, the RSD was substantially reduced. Gender-specific differences in the RSD of the absolute volumes were found in the gyrus rectus and the orbital gyri. These differences were only partially compensated for by volume normalization. The cerebral gyral segments also showed a general trend for decreasing volumes with increasing age, but there was no significant correlation (*Supplemental Figure 4*).

**Figure 2.**
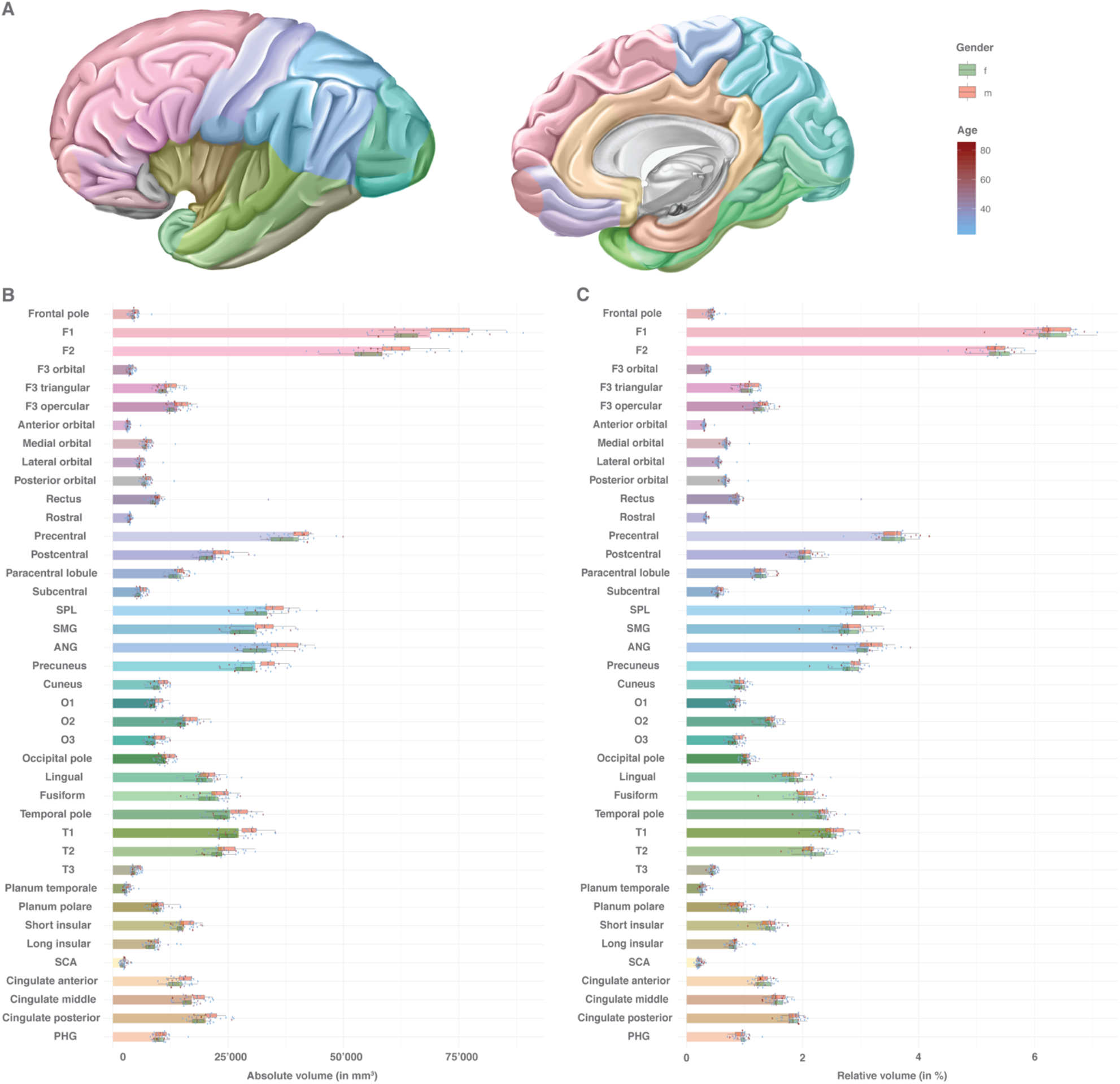
Volumes of the cerebral gyral segments: Absolute and relative volumes of the cerebral gyral segments. **A:** Color scheme of the anatomy, gender and age. **B:** Absolute volumes in mm^3^. Mean volumes are given as bars. Gender stratification is provided with boxplots. The individual measurements are shown as dots with continuous color-coding indicating age. **C:** Relative volumes normalized to the total individual encephalic volume (without ventricles). The graphic elements follow B. *Abbreviations*: ANG = angular gyrus; f = female; F1 = superior frontal gyrus; F2 = middle frontal gyrus; F3 = inferior frontal gyrus; m = male; O1 = superior occipital gyrus; O2 = middle occipital gyrus; O3 = inferior occipital gyrus; PHG = parahippocampal gyrus; SCA = subcallosal area; SMG = supramarginal gyrus; SPL = superior parietal lobule; T1 = superior temporal gyrus; T2 = middle temporal gyrus; T3 = inferior temporal gyrus.

### Central prosencephalic structures

The volumes of the subgyral central prosencephalic structures are reported in *Supplemental Table 4* and *Figure 3*. The thalamus represents the most voluminous central prosencephalic structure (1.3%, SD 0.10, RSD 7.6%). Among the basal ganglia, 51% of the volume belongs to the putamen, 35% to the caudate nucleus and 14% to the globus pallidum. While all the other structures were more voluminous in males than in females, the volume of the corpus callosum was higher in females. There was some evidence for a gender difference in the relative volumes of the thalamus (p = 0.016) and the corpus callosum (p = 0.033), while a normalization to the total individual encephalic volume resulted in comparable relative volumes for all other structures. The RSD was higher in smaller anatomical structures (e.g. 31.1% in the claustrum, 19.3% in the globus pallidum) but without any relevant gender effect. Normalization to the total individual encephalic volume generally led to a mild reduction in relative variance. The corpus callosum, thalamus, putamen, hippocampus and amygdala showed a significant reduction in absolute volumes with increasing age (*Supplemental Figure 5*). This effect was moderately counteracted by normalization to the total individual encephalic volume. Other anatomical structures, such as the claustrum, the internal capsule, the caudate and the globus pallidum, as well as the hypothalamus, demonstrated a tendency for decreasing volumes with increasing age, but there was only weak to no evidence for a correlation.

**Figure 3.**
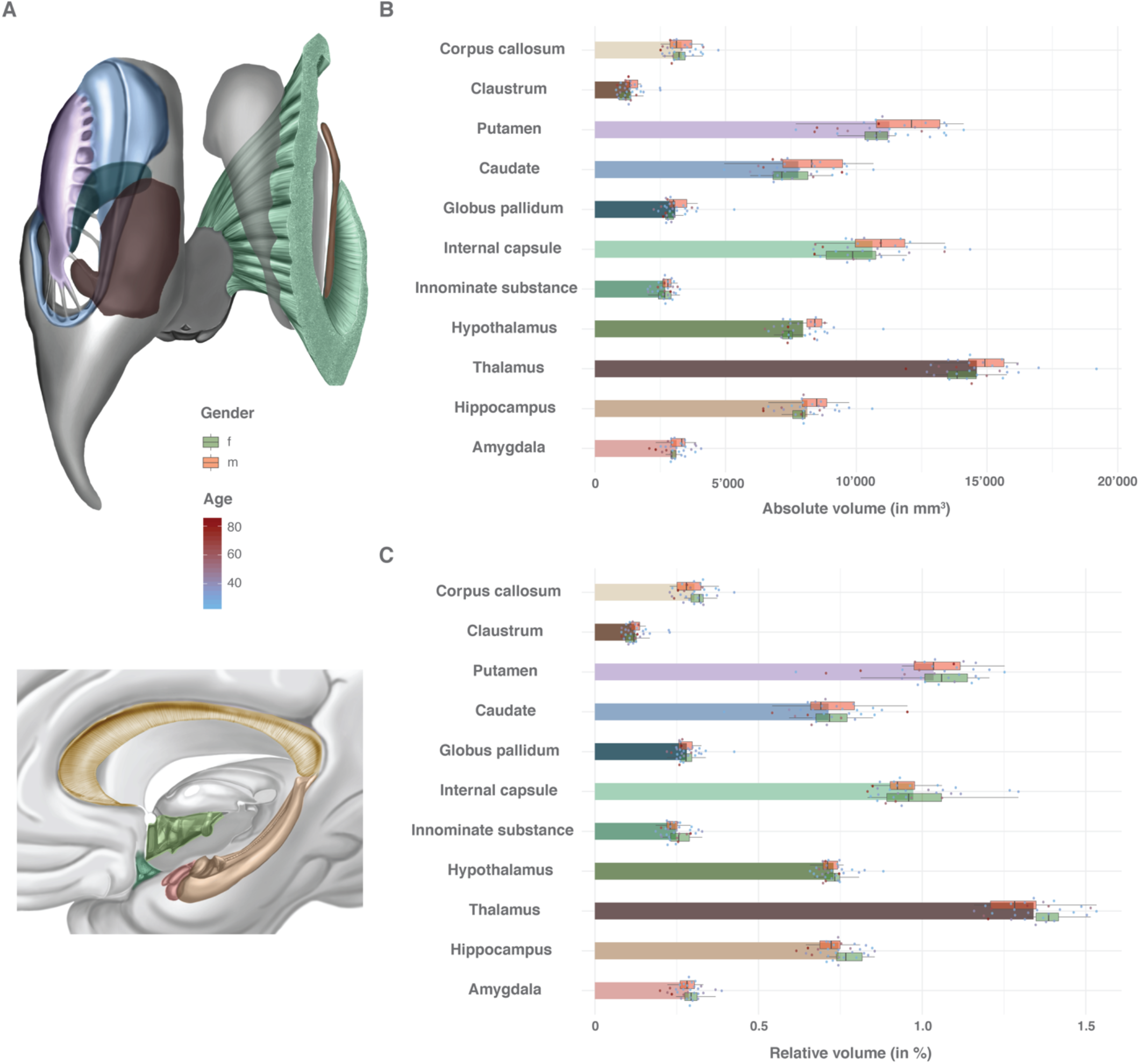
Volumes of central prosencephalic structures: Absolute and relative volumes of the anatomical white and gray matter structures of the central prosencephalon. **A:** Color scheme of the anatomy, gender and age. **B:** Absolute volumes in mm^3^. Mean volumes are given as bars. Gender stratification is provided with boxplots. The individual measurements are shown as dots with continuous color-coding indicating age. **C:** Relative volumes normalized to the total individual encephalic volume (without ventricles). The graphic elements follow B. *Abbreviations*: f = female; m = male.

### Brainstem and cerebellum

*Supplemental Table 5* and *Figure 4* give the volumes of the various structures of the brainstem and the cerebellum. The pons makes up 54% of the brainstem, 36% is made up of the mesencephalon and only 10% is made up of the medulla oblongata. The vermis accounts for 5% of the total cerebellum. The posterior lobe dominates with 47% of the cerebellar volume, followed by the anterior and medial lobe with comparable relative volumes (28% and 25%, respectively). The flocculonodular lobe is the smallest one with a relative cerebellar volume of 1%. *Supplemental Table 5* and *Figure 4D&E* also provide the volumes of the individual cerebellar vermal and hemispheric lobules. The most voluminous vermal lobule is the Culmen, while the largest hemispheric lobule is the inferior semilunar/gracile lobule. In males, there was a tendency for larger volumes for all lobules except the vermian central lobule and the hemispheric superior semilunar lobule. This tendency was balanced by the normalization to total individual encephalic volume. The relative variance between the infratentorial structures showed comparable results. The small cerebellar lobular structures had a slightly increased RSD. No distinct difference in RSD was found between genders. The normalization to the total individual encephalic volume had only a mild variance-reducing effect. While the pons and cerebellar peduncles demonstrated an increasing volume with age, the other structures tended to decrease in volume with increasing age. This effect was most pronounced in the medial cerebellar lobe (*Supplemental Figure 6*).

**Figure 4.**
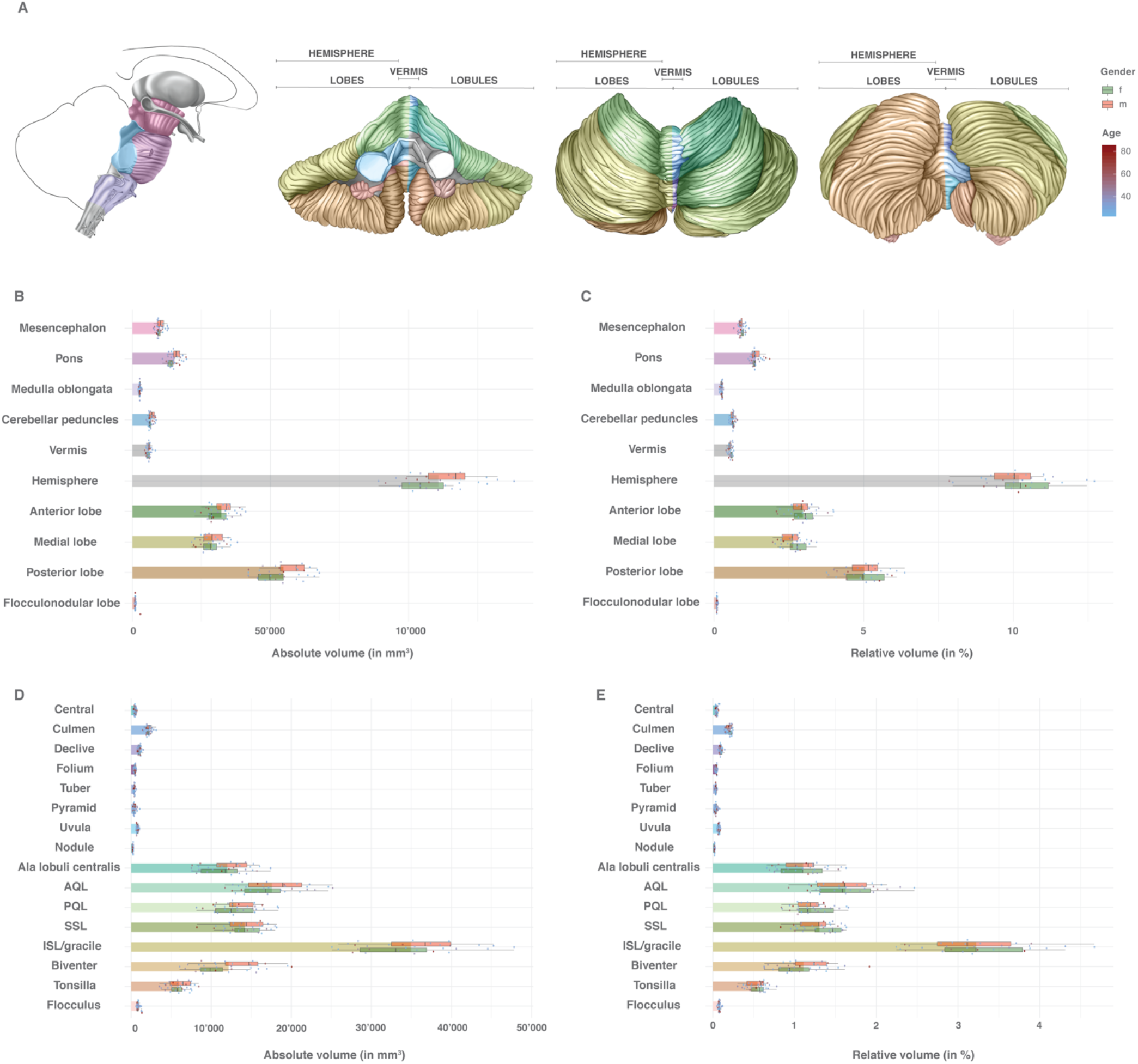
Volumes of brainstem and cerebellar structures: Absolute and relative volumes of the anatomical structures of the brainstem and the cerebellum. **A:** Color scheme of the anatomy, gender and age. **B:** Absolute volumes of the mesencephalon, pons, medulla oblongata as well as cerebellar peduncles and cerebellar lobes in mm^3^. Mean volumes are given as bars. Gender stratification is provided with boxplots. The individual measurements are shown as dots with continuous color-coding indicating age. **C:** Relative volumes of the mesencephalon, pons, medulla oblongata as well as cerebellar peduncles and cerebellar lobes. Volumes were normalized to the total individual encephalic volume (without ventricles). The graphic elements follow B. **D:** Absolute volumes of the cerebellar vermian and hemispheric lobules in mm^3^. The graphic elements follow B. **E:** Relative volumes of the cerebellar vermian and hemispheric lobules normalized to the total individual encephalic volume (without ventricles). The graphic elements follow B. *Abbreviations*: f = female; m = male.

### Ventricular system

The ventricular system has a total median volume of 21 185 mm^3^ (SD 16 714, range 7987 to 71 384) with a median relative volume (to the total individual encephalon without ventricle) of 1.93% (SD 1.52, RSD 78.7%). Details are given in *Supplemental Table 6* and *Figure 5*. The lateral ventricles occupy 83.2% (SD 6.84, RSD 8.2%) of the ventricular system, the third ventricle 6.0% (SD 1.79, RSD 29.7%) and the fourth ventricle 10.8% (SD 5.59, RSD 51.9%). Within the lateral ventricles, the frontal horn (28.1%, SD 4.70, RSD 16.7%) dominates in volume, followed by the atrium (24.2%, SD 5.96, RSD 24.6%) and the body (21.1%, SD 4.93, RSD 23.3%). The temporal horn is least prominent (4.8%, SD 2.74, RSD 56.7%), close to the occipital horn (4.9%, SD 2.61, RSD 52.6%). Within the fourth ventricle, the apex (1.1%, SD 0.67, RSD 60.7%), the lateral recess (1.3%, SD 0.66, RSD 50.2%), the obex (1.1%, SD 0.85, RSD 74.2%) and the fastigium (1.3%, SD 0.66, RSD 50.4%) have comparable relative volumes. The absolute volumes tended to be larger in males than in females. Both the normalization to the total individual encephalic volume and to the total individual ventricular volume compensated for this effect. The total ventricular system, as well as its subdivisions, demonstrate a substantially higher variance than the abovementioned parenchymal structures, as evidenced by the RSD values. The relative variance decreases from the lateral, through the third, to the fourth ventricle. Within the lateral ventricle, the temporal horn exhibits the smallest variance. No gender-specific differences in relative variance were identified. A normalization to the total individual encephalic volume did not result in a relevant reduction in the relative variance of the ventricular system and its divisions. By a normalization to the total individual ventricular volume, however, RSD values approaching those of the parenchymal structures were achieved. Ventricular volumes showed a significant and pronounced increase with age (*Supplemental Figure 7*). While a normalization to the total individual encephalic volume further enhances this effect, it is compensated for by a normalization to the total individual ventricular volume.

**Figure 5.**
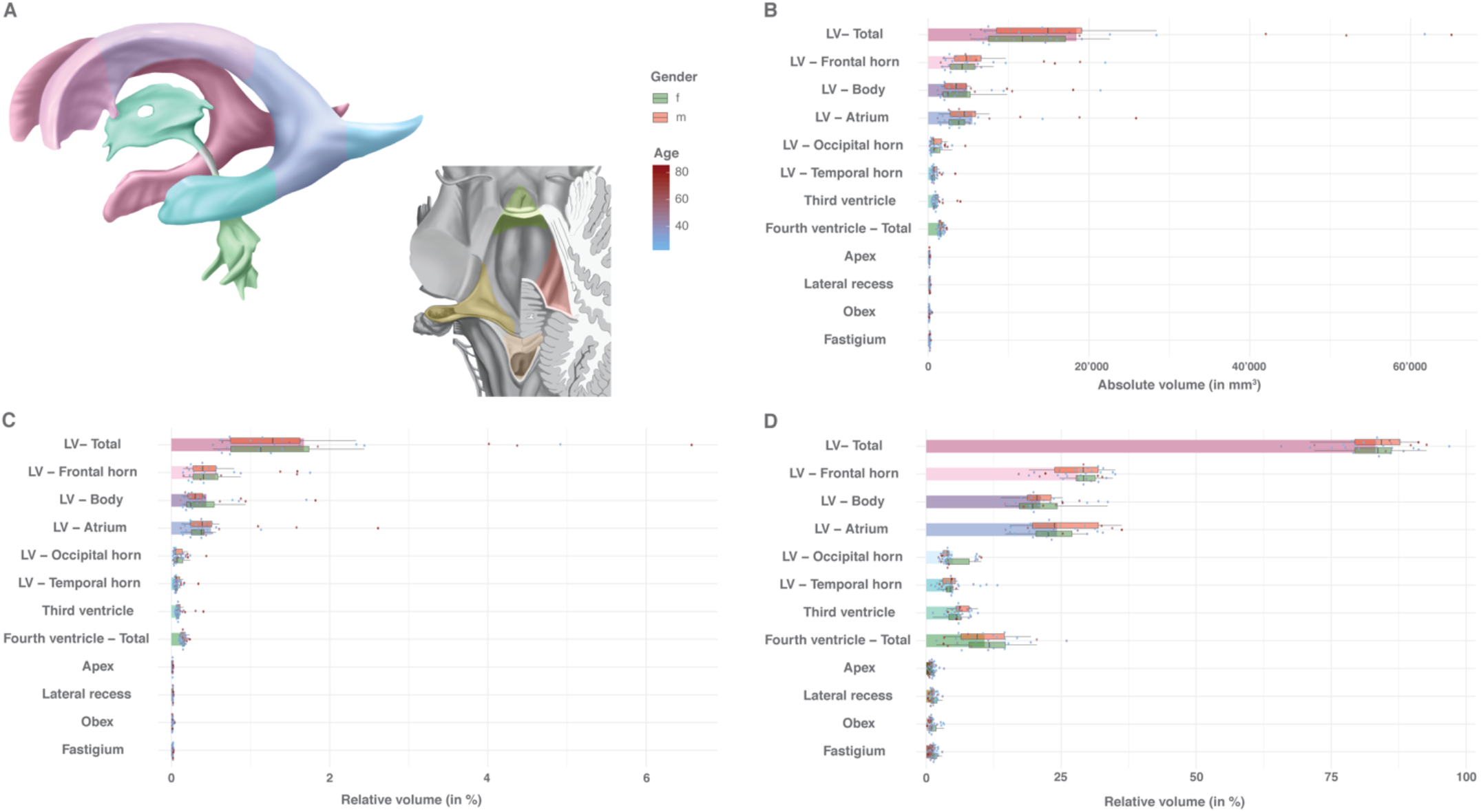
Volumes of the ventricular system: Absolute and relative volumes of the anatomical divisions of the ventricular system. **A:** Color scheme of the anatomy, gender and age. **B:** Absolute volumes in mm^3^. Mean volumes are given as bars. Gender stratification is provided with boxplots. The individual measurements are shown as dots with continuous color-coding indicating age. **C:** Relative volumes normalized to the total individual encephalic volume (without ventricles). The graphic elements follow B. **D:** Relative volumes normalized to the total individual ventricular volume. The graphic elements follow B. *Abbreviations*: f = female; LV = lateral ventricle; m = male.

### Reference dataset

Complete data on the volumes of every parcellation unit for each subject and side are provided in the *Supplemental Raw Data*.

## Discussion

The current investigation provides an anatomically detailed and comprehensive analysis of the absolute and relative volumes of the human encephalic structures using a clinically oriented parcellation algorithm. This reference is intended to serve for volume-standardization in clinical studies on the topographic prevalence of pathologies. In addition, the supplied parcellation algorithm and dataset provides the basis for a reference database for analyses of the variability of these structures.

### The novelty of the current volume-standardization atlas in comparison to existing brain atlases

Quantitative assessments of brain size and its variations, relating to evolution, gender, age or pathology, feature prominently in neuroscience (Harper and Mina 1981; Stephan et al. 1988; Coffey et al. 1992; Finlay and Darlington 1995; Barton and Harvey 2000; Clark et al. 2001; Good et al. 2001). In comparison to other organs, special demands on morphometry arise from the anatomical complexity of the human brain. Classic pathological analyses, despite their high structural sensitivity, are subject to limitations caused by fixation and postmortem tissue changes, which apply, in particular, to the structures of the brain (Filipek et al. 1989; Good et al. 2001). These limitations can nowadays be overcome by morphometric analyses based on in-vivo neuroimaging. While the study of *Filipek et al*. (Filipek et al. 1994) focused exclusively on larger anatomical areas, *Kennedy et al*. (Kennedy 1998) and *Crespo-Facorro et al*. (Crespo-Facorro et al. 2000) provided more detailed volumes of the cerebral cortical structures. *Chiavaras et al*. (Chiavaras et al. 2001) further characterized the orbitofrontal sulci and *Hammers et al*. (Hammers et al. 2003) focused on the temporal lobe. *Klein et al*. (Klein et al. 2005), *Heckemann et al*. (Heckemann et al. 2006) and *Shattuck et al*. (Shattuck et al. 2008) provided more volumetric details on the cortical gyral areas. The study presented here complements these analyses and provides an anatomically detailed and comprehensive reference on the relative volumes of encephalic structures considered relevant for studies on the topographic prevalence of brain pathologies.

### Absolute volumes

The reported mean total encephalic volume of 1 093 437 mm^3^ (SD 111 353) is within the range of previous studies, for example that of *Filipek et al*. (Filipek et al. 1994) with 1 380 100 mm^3^ (SD 113 900), *Hammers et al*. (Hammers et al. 2003) with 1 289 861 mm^3^ (SD 147 973) or *Lancaster et al*. (Lancaster et al. 2010) with 1 179 978 mm^3^ (SD 112 537). The absolute lobar volumes are slightly below those in the aforementioned studies, most likely due to differences in the lobar classification systems used. The reported volumes of the gyral segments are higher than the volumes described by *Shattuck et al*. (Shattuck et al. 2008), *Hammers et al*. (Hammers et al. 2003), *Crespo-Facorro et al*. (Crespo-Facorro et al. 2000) or *Kennedy et al*. (Kennedy et al. 1998). This is best explained in that our study analyzed the entire gyral segment (cortex and white matter), while the aforementioned studies only considered the cortical volume of a gyral segment. Isolated cortical volumes are provided in the *Supplemental Raw Data* for our study. Since neoplastic or vascular pathologies are often not only localized cortically, but rather affect whole gyral segments (Yasargil 1994; Akeret et al. 2020), the approach adopted in this study is tailored to volume-standardization in topographic studies. This study provides one of the most detailed volumetric analyses of the central prosencephalic anatomy, infratentorial structures and anatomical segments of the ventricular system. The volumes of the corpus callosum (Hammers et al. 2003), the putamen (Filipek et al. 1994; Hammers et al. 2003; Shattuck et al. 2008; Lancaster et al. 2010), the caudate (Filipek et al. 1994; Hammers et al. 2003; Shattuck et al. 2008; Lancaster et al. 2010), the globus pallidum (Filipek et al. 1994; Hammers et al. 2003), the thalamus (Hammers et al. 2003), the hippocampus (Filipek et al. 1994; Hammers et al. 2003; Shattuck et al. 2008; Lancaster et al. 2010) as well as the amygdala (Filipek et al. 1994; Hammers et al. 2003) are comparable to previous reports. The volumes reported for the brainstem (Filipek et al. 1994; Hammers et al. 2003; Shattuck et al. 2008; Lancaster et al. 2010) and the cerebellum (Filipek et al. 1994; Hammers et al. 2003; Shattuck et al. 2008; Lancaster et al. 2010) are consistent with previous studies. The cerebellum has been claimed to represent a phylogenetically-preserved constant fraction of the total encephalic volume of 0.13 (SD 0.02) across mammalian taxa (Clark et al. 2001), an approximation consistent with our results (10.7%, SD 1.17). The total ventricular volume (DeLisi et al. 1991; Shenton et al. 1992b; Coffey et al. 1992; Filipek et al. 1994) as well as the volumes of the lateral ventricles (Filipek et al. 1994), the third (Filipek et al. 1994; Hammers et al. 2003) and the fourth ventricle (Filipek et al. 1994) correspond to previous reports.

### Volume variability and normalization

Consistent with previous studies (Filipek et al. 1994; Lancaster et al. 2010), we report a considerable interindividual variability in the volumes of neuroanatomical structures. This variability renders comparative analyses difficult. A normalization accounting for interindividual natural variance as well as age- and gender-specific differences would facilitate such comparisons. *Lancaster et al*. (Lancaster et al. 2010) described that a normalization to the entire cerebral hemisphere best counteracts variance, but the study lacked anatomical detail. In the present study, this concept is expanded to the entire encephalon with greater anatomical depth. A normalization to the total individual encephalon reduced the relative variance substantially for most parenchymal structures. The ventricular structures, however, maintained a high variance after normalization to the total individual encephalic volume. A normalization of the divisions of the ventricular system to the total individual ventricular volume, however, reduced the interindividual variability more accurately. The most likely explanation lies in a differing dependence of the brain parenchyma and the ventricular system on age, as described below. No gradient corresponding to a phylo-or ontogenetic pattern was identified in the comparison of the relative volumetric structural variance. Female brains have been reported to be approximately 10% smaller than male brains (Zatz et al. 1982; Swaab and Hofman 1984; Hammers et al. 2003). This roughly corresponds to the gender difference in the absolute volumes of our study for the total encephalic volume (12.5%, 1 024 921.71 mm^3^ vs. 1 153 388.38 mm^3^), but also for the individual anatomical structures (*Tables 1-5, Figures 1-5*). For almost all structures, this gender effect was compensated for by normalization. Only the corpus callosum, the thalamus, the medial cerebellar lobe as well as the central vermian and the superior semilunar lobule yielded evidence of a gender difference after normalization. Given the context of multiple testing, this evidence should be interpreted with caution. A gradual decrease in brain volume with increasing age, beginning after the fourth to fifth decade of life, has been described, with a variable rate of between 1% and 3.5% per decade (Dekaban and Sadowsky 1978; Miller et al. 1980; Harper and Mina 1981; Hubbard and Anderson 1981; Hatazawa et al. 1982). Although our study did not show a significant correlation between the age of the subjects and the volume of most parenchymal structures, it revealed a general downward trend with increasing age. The absence of significance might be due to a limited power given the number of subjects and the fact that most of them were under 50 years of age. The direction and magnitude of the observed trend nevertheless correspond to previous observations. In contrast to the brain parenchyma, a significant increase in volume was observed with increasing age for the ventricular system. This inverse trend between brain parenchyma and ventricular system supports a normalization of the ventricular segments to its total volume rather than to the total encephalic volume.

### Limitations and strengths

First, due to scan time limitation, the resolution of the T1-weighted scan was about 0.64 mm^3^. According to *Aldusary et al*. (*Supplemental Figure 5* of the *Supplemental Data*)(Aldusary et al. 2019), the data resolution dependent uncertainty of volume estimates can be asserted based on the simulated discretization noise. For the smallest volumes estimated in this study (apex (163 mm^3^), lateral recess (199 mm^3^), obex (174 mm^3^), and fastigium (199 mm^3^) of the fourth ventricle), the discretization noise has a standard deviation of approximatively 2 mm^3^. On the other hand, segmentation of the lingula and vinculum of the cerebellum was omitted due to their small volume and the associated volumetric inaccuracy. Second, the hybrid methodology combining both manual and semi-automated segmentation might be a potential limitation. While certain anatomical delineations are highly consistent and easily automated, others – especially at a detailed anatomical level – are dependent on expert judgement (Hammers et al. 2003). More resource-efficient semi-automated segmentation was used for those structures for which reliable protocols were available. The remaining structures were supplemented by manual segmentation to achieve the highest possible accuracy (Crespo-Facorro et al. 2000; Shattuck et al. 2008). The multi-layer parcellation allowed additional plausibility checks, in which the volumes of larger structures (e.g. the brainstem as a whole in the semi-automated analysis) were verified against the sum of their components (mesencephalon, pons and medulla oblongata in manual analysis). Thus, overall, the risk of bias from this hybrid approach is considered to be small. Third, the adopted cerebral lobar classification is unconventional. It distinguishes a limbic and a central lobe (Yasargil 1994), representing structural, functional, and pathoclitic units (Yasargil 1994; Akeret et al. 2019, 2020). Owing to the multi-level parcellation method, however, it is also possible to determine the absolute and relative volumes of the lobes corresponding to other lobar classification systems based on the individual gyral segmental volumes. The same principle is applied for alternative cerebellar lobar classifications. Fourth, due to the high number of parcellation units and the limited number of subjects, the conclusions regarding gender- and age-effects are limited. The aim of this study was not to draw definite conclusions on volume-variability, but to serve as a reference for relative volumes in studies on the topographic prevalence of brain pathologies. In addition, it may provide the basis for a future reference dataset to analyze volume variability. Given the multiple testing, a strict significance level was not defined, but rather the strength of evidence for a difference was provided. Due to the same limitations regarding statistical power, bilateral volumetric symmetry was not analyzed in this study, nor was the possible influence of handedness on such. In bilateral structures, the volumes of the left- and the right-side structures were combined to control the total number of variables. The lack of an assessment of the interrater reliability constitutes another limitation in our analysis. Finally, no automated parcellation tool is provided in the current study, as a such does not assist in the volume-normalization of disease topography. However, a such may be useful in the future to assess the absolute and relative variability of brain structures in a larger cohort.

### Practical implications

Clinical studies on the topography of pathologies are often limited by the absence of a normalization of the prevalence of pathologies to the relative volume of the affected anatomical structures. The present study aims to provide an anatomically detailed and comprehensive atlas for topographic volume-standardization in pathoclitic studies. Identifying and characterizing such topographic patterns may provide important pathophysiological insights, inform diagnostic criteria and pave the way for anatomically targeted therapeutic approaches. In addition, the neuroanatomical parcellation algorithm and the dataset supplied form the basis for establishing a reference population of clinically relevant absolute and relative neuroanatomical volumes and for the analyses of their variability.

## Conclusions

The present study constitutes an anatomically detailed and comprehensive analysis of the absolute and relative volumes of the human encephalic structures using a clinically oriented parcellation algorithm. This reference is intended to serve for volume-standardization in clinical studies on the topographic prevalence of pathologies. In addition, the supplied parcellation algorithm and dataset provides the basis for a reference database for analyses of the variability of these structures.

## Data Availability

The authors confirm that the data supporting the findings of this study are available within the article and its supplemental materials. In addition, the anonymized complete dataset is provided as a basis for further investigations in the Supplemental Raw Data.

## Acknowledgments

Kevin Akeret is supported by the Prof. Dr. med. Karl und Rena Theiler-Haag foundation (F-86101-26-01). Jorn Fierstra is supported by the Swiss Cancer League (KFS 3975-08-2016-R). We thank Miller Rasmy for his contribution to the artistic work.

## Abbreviations

MPRAGE: Magnetization Prepared Rapid Acquisition Gradient Echo
ROI: region of interest
RSD: relative standard deviation
SD: standard deviation

## Declarations

### Ethical approval

Ethics board approval was obtained prior to data collection (KEK ZH 2012-0427).

### Consent to participate/Consent for publication

All patients or their legal representatives gave their written informed consent for participation and publication.

### Availability of data and material

Additional data is made available in the Supplemental Information online, including the source dataset.

### Code availability

The statistical software (R) code used is made available in the supplementary methods online.

### Author’s contributions

Kevin Akeret: Concept and design; Acquisition of data; Analysis and interpretation of data; Drafting the article; Statistical analysis; Reviewed submitted version of manuscript.

Christiaan Hendrik Bas van Niftrik: Acquisition of data; Drafting the article; Reviewed submitted version of manuscript.

Martina Sebök: Acquisition of data; Reviewed submitted version of manuscript.

Giovanni Muscas: Acquisition of data; Reviewed submitted version of manuscript.

Thomas Visser: Acquisition of data; Reviewed submitted version of manuscript.

Victor E. Staartjes: Concept and design; Reviewed submitted version of manuscript.

Federica Marinoni: Acquisition of data; Reviewed submitted version of manuscript.

Carlo Serra: Concept and design; Reviewed submitted version of manuscript.

Luca Regli: Concept and design; Reviewed submitted version of manuscript.

Niklaus Krayenbühl: Concept and design; Administrative/technical/material support; Reviewed submitted version of manuscript.

Marco Piccirelli: Drafting the article; Reviewed submitted version of manuscript.

Jorn Fierstra: Concept and design; Drafting the article; Administrative/technical/material support; Study supervision; Reviewed submitted version of manuscript.

