## Supplemental Material for "Topographic volume-standardization atlas of the human brain"

#### Supplemental Figures

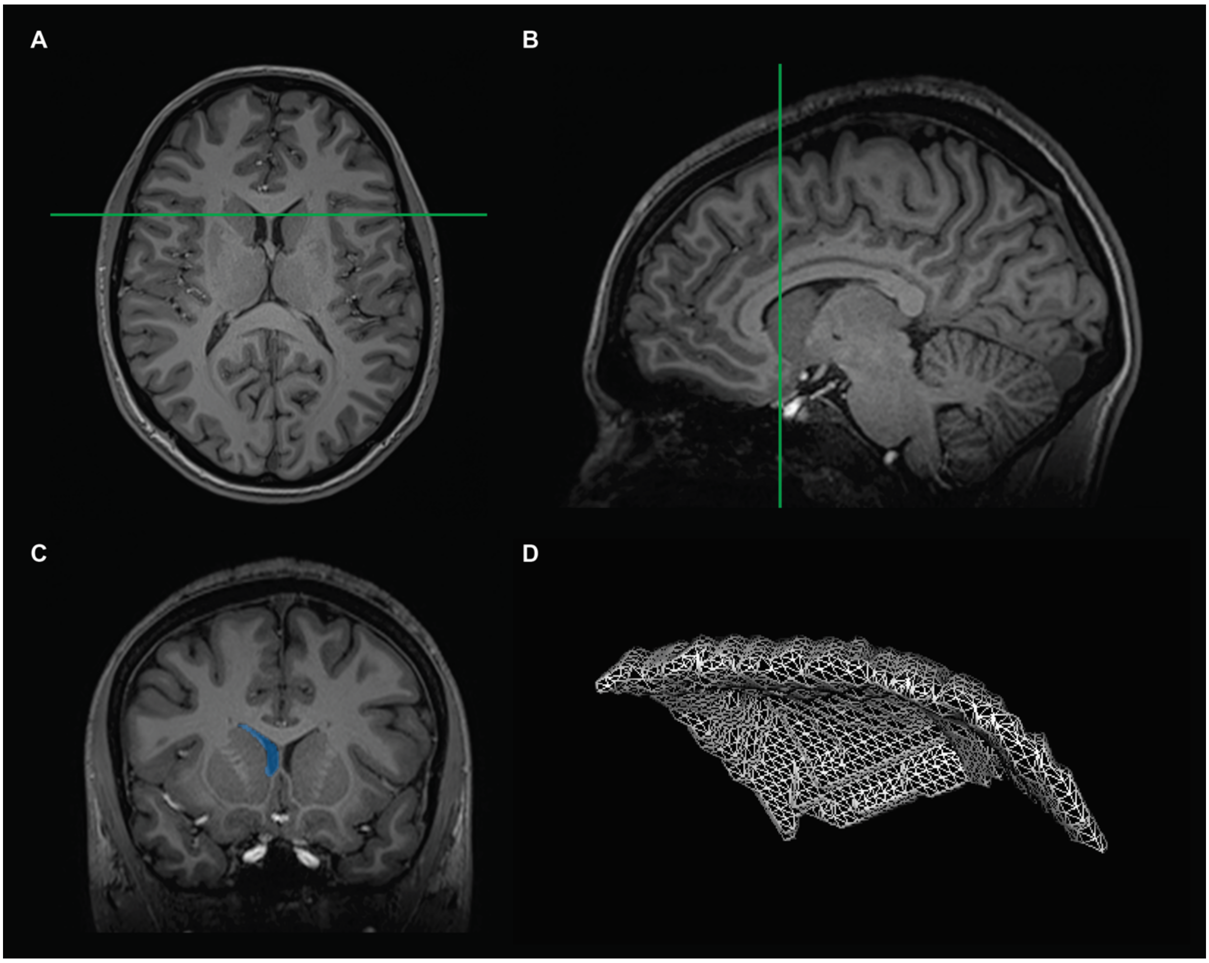


**Supplemental Figure 1. Manual segmentation:** The manual anatomical outline of a parcellation unit is exemplified on the right frontal horn of subject 19. **A:** Axial plane with the coronal slice indicated. **B:** Sagittal plane with the coronal slice indicated. **C:** Coronal plane with the segmentation of the right frontal horn shown in blue. **D:** Three-dimensional reconstruction of the right frontal horn from segmentation in all slices guided by the neuroanatomical parcellation algorithm of *Supplemental Table 1*.

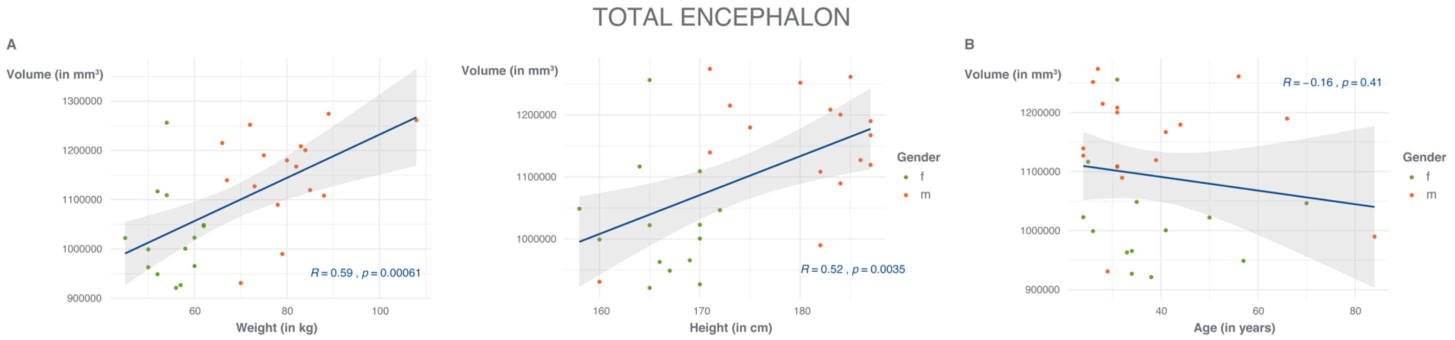


**Supplemental Figure 2. Total encephalic volume: A:** Absolute total encephalic volume correlated to the weight (left) and the height (right) of the subjects and stratified by gender. *Abbreviations*: f = female; m = male. **B:** Absolute total encephalic volume correlated to age and stratified by gender.

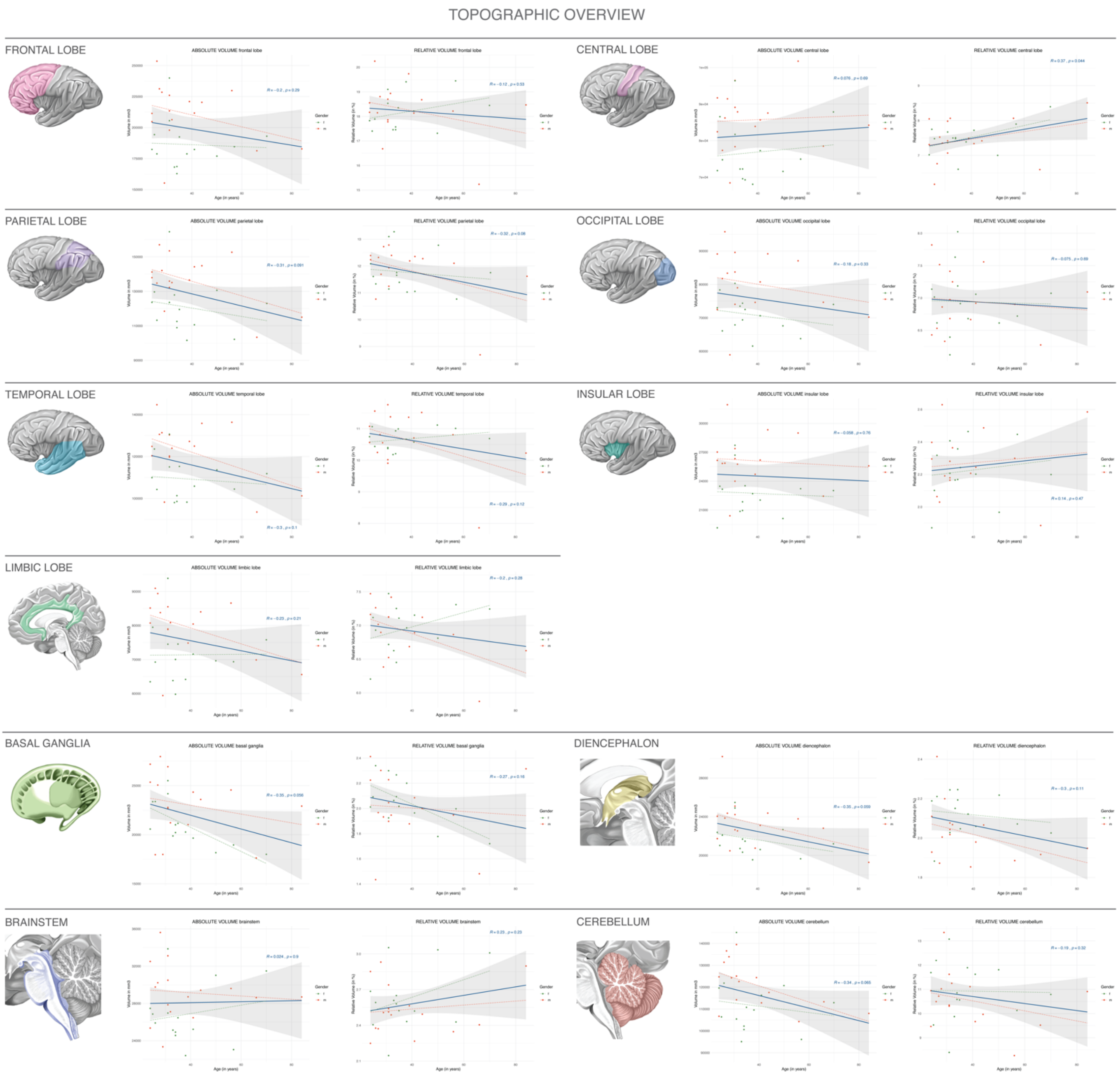


**Supplemental Figure 3. Volumes of general anatomical structures correlated to age:** Absolute and relative volumes of the cerebral lobes, basal ganglia, diencephalon, brainstem and cerebellum in correlation to the age of the subjects. The specific anatomical structures are illustrated schematically, the corresponding left diagram displays the correlation between absolute volume and age, the right diagram the correlation between relative volume and age. Volumes were normalized to the total individual encephalic volume (without ventricles). Color-coding provides a stratification by gender. *Abbreviations*: f = female; m = male.

### CEREBRAL GYRI

#### GYRI OF THE FRONTAL LOBE

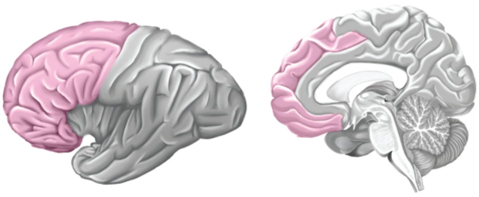

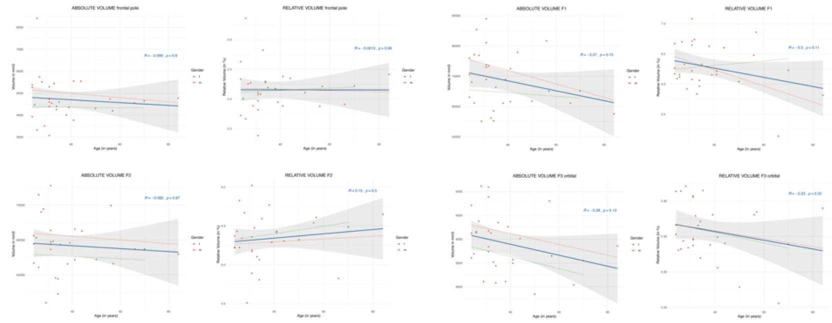

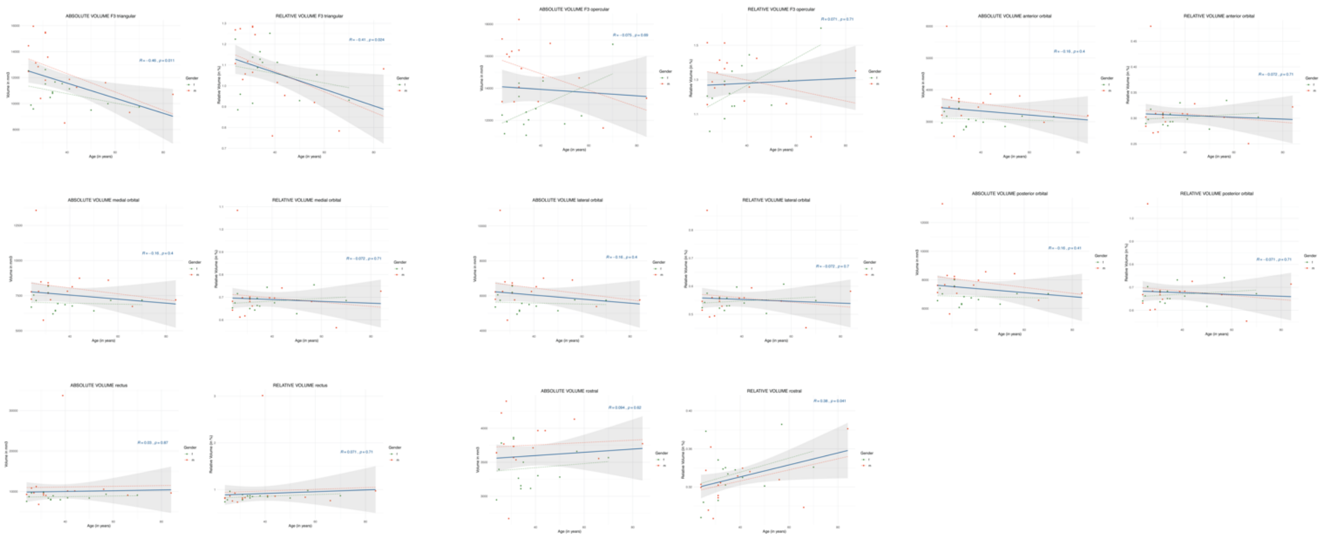

#### GYRI OF THE CENTRAL LOBE

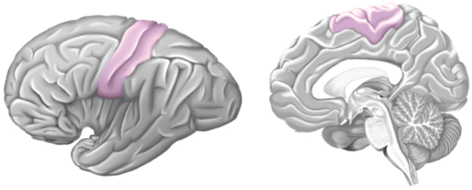

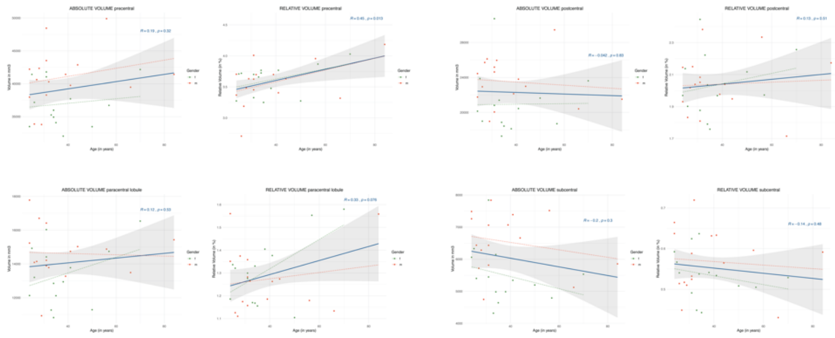

#### GYRI OF THE PARIETAL LOBE

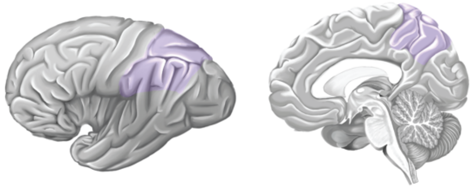

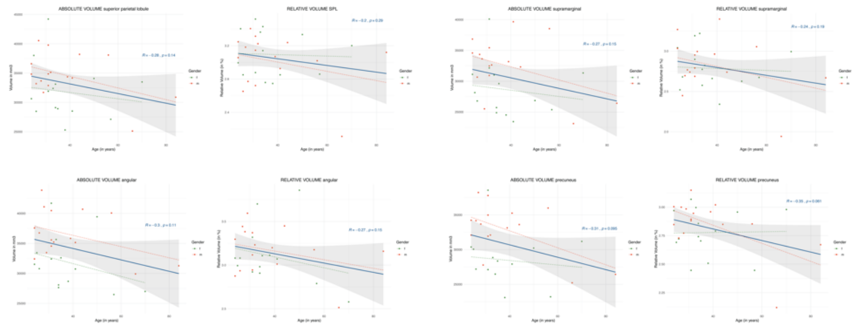

#### GYRI OF THE OCCIPITAL LOBE

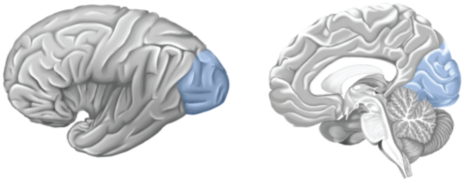

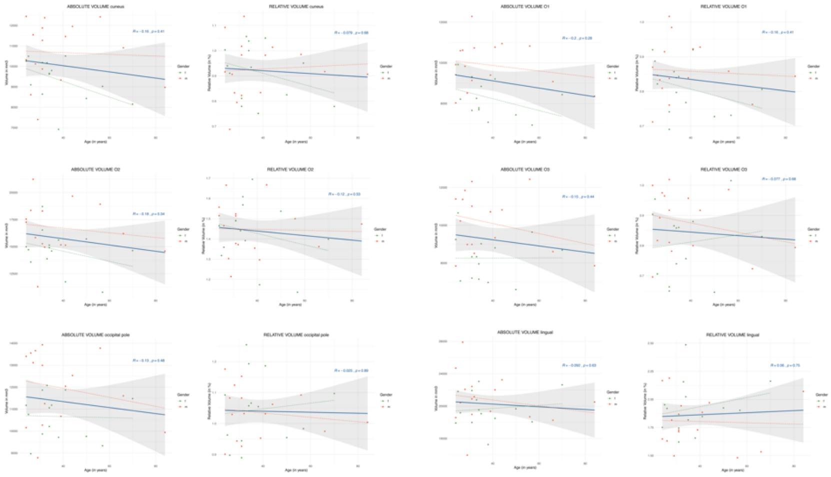


#### GYRI OF THE TEMPORAL LOBE

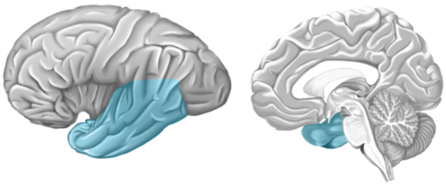

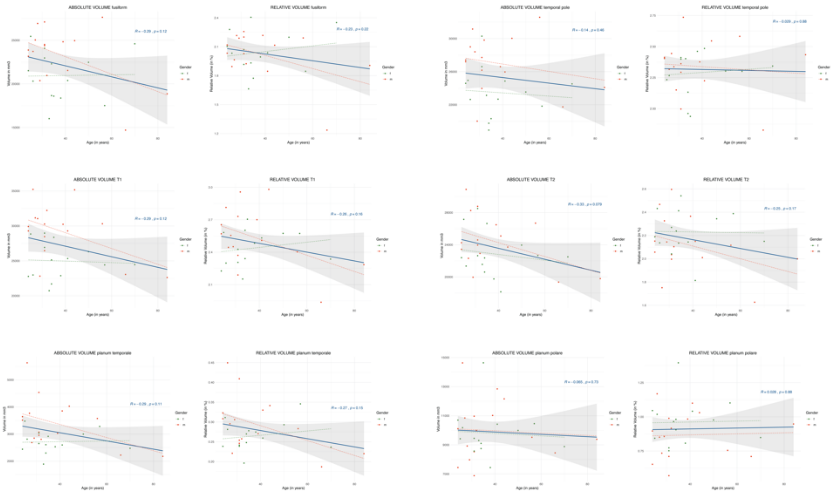


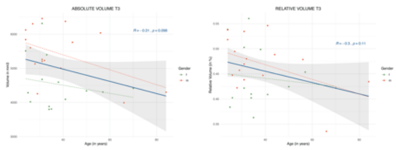


#### GYRI OF THE INSULAR LOBE

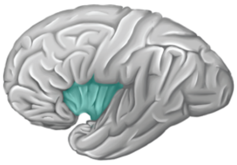

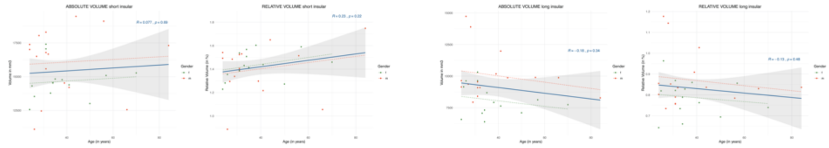


#### GYRI OF THE LIMBIC LOBE

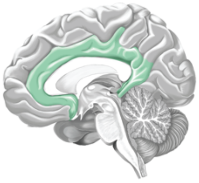

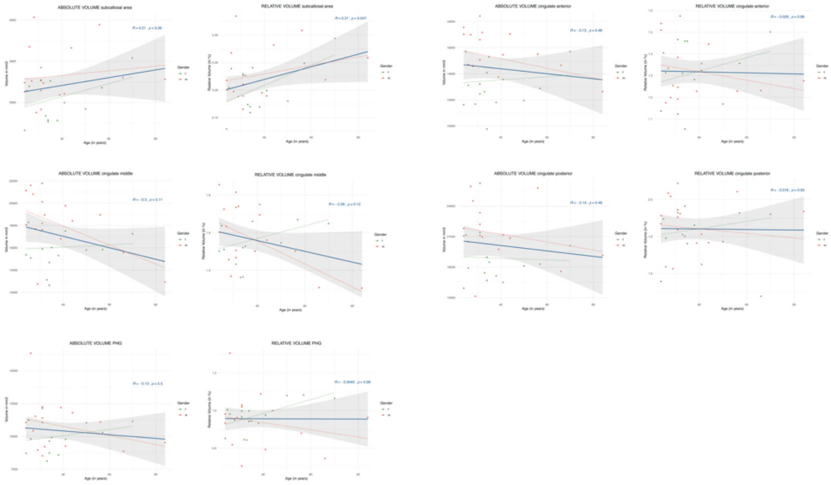

**Supplemental Figure 4. Volumes of cerebral gyral segments correlated to age:** Absolute and relative volumes of the cerebral gyral segments in correlation to the age of the subjects. The gyral segments are grouped according to the corresponding cerebral lobes, as illustrated schematically. For each gyral segment, the left diagram gives the correlation between absolute volume and age, the right diagram the correlation between relative volume and age. Volumes were normalized to the total individual encephalic volume (without ventricles). Color-coding provides a stratification by gender. *Abbreviations*: f = female; m = male.

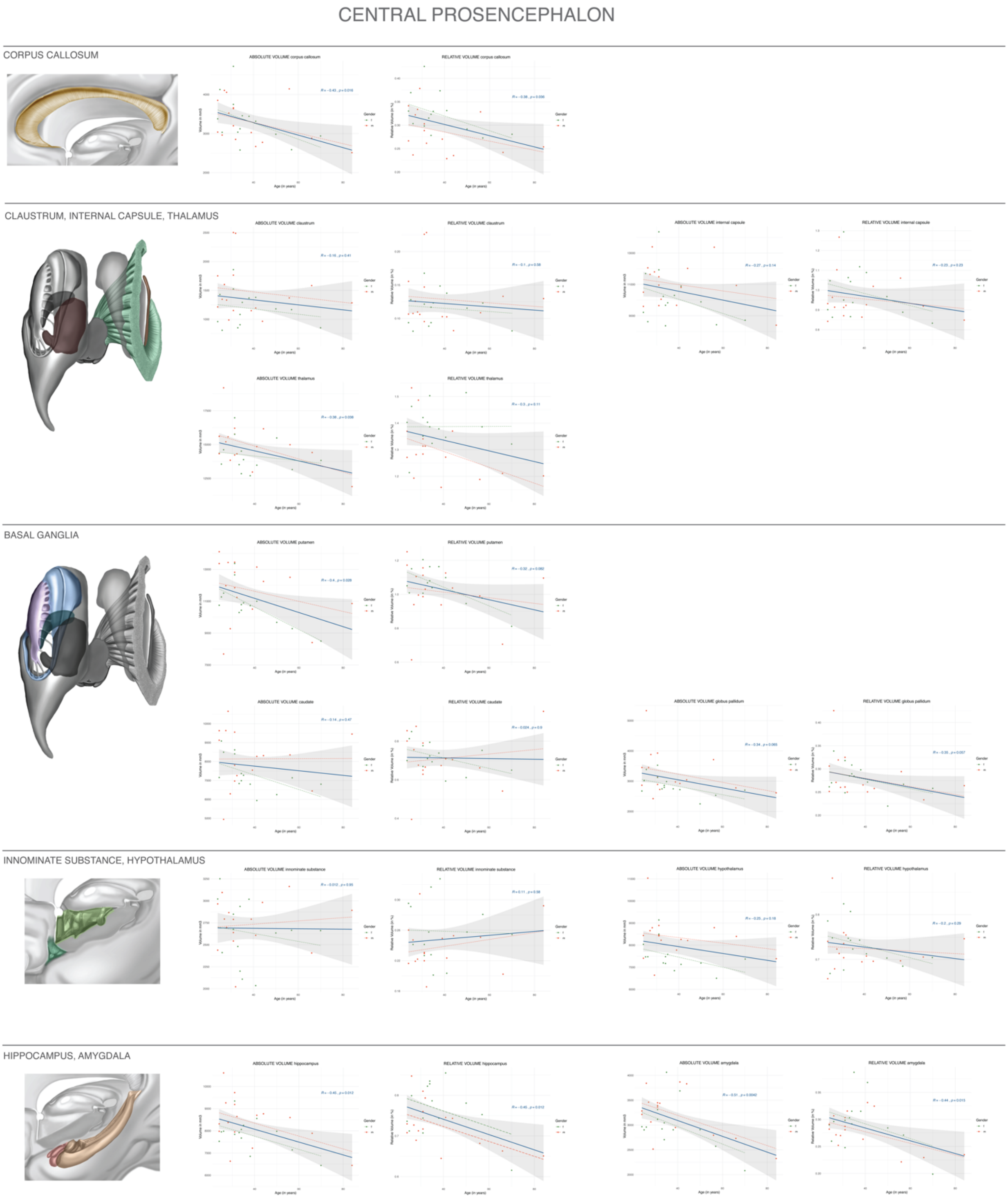


**Supplemental Figure 5. Volumes of central prosencephalic structures correlated to age:** Absolute and relative volumes of the central prosencephalic white and gray matter structures in correlation to the age of the subjects. The specific anatomical structures are illustrated schematically. For each structure, the left diagram displays the correlation between absolute volume and age, the right diagram the correlation between relative volume and age. Volumes were normalized to the total individual encephalic volume (without ventricles). Color-coding provides a stratification by gender. *Abbreviations*: f = female; m = male.

### BRAINSTEM & CEREBELLUM

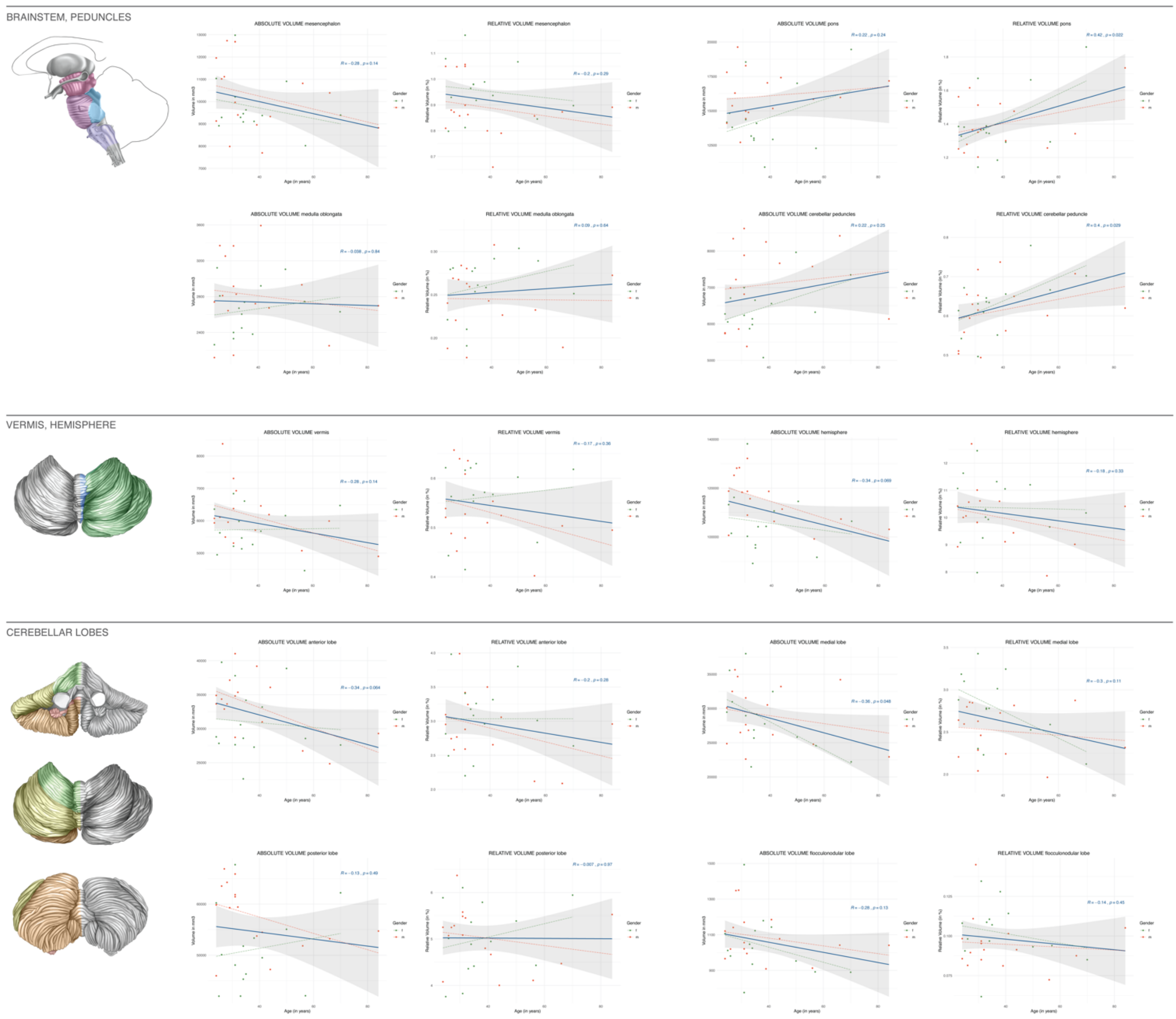


**Supplemental Figures 6. Volumes of brainstem and cerebellar structures correlated to age:** Absolute and relative volumes of the brainstem, the cerebellar vermis and hemisphere as well as the cerebellar lobes in correlation to the age of the subjects. The specific anatomical structures are illustrated schematically. For each structure, the left diagram displays the correlation between absolute volume and age, the right diagram the correlation between relative volume and age. Volumes were normalized to the total individual encephalic volume (without ventricles). Color-coding provides a stratification by gender. *Abbreviations*: f = female; m = male.

### VENTRICULAR SYSTEM

#### TOTAL VENTRICULAR SYSTEM

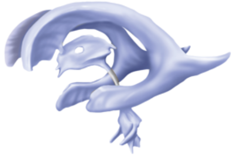

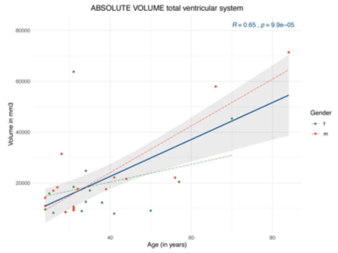


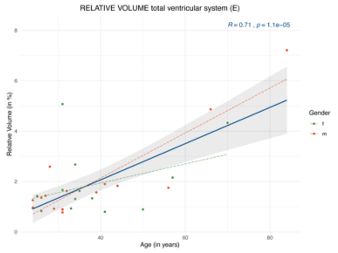


#### LATERAL VENTRICLE

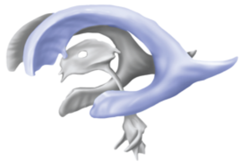

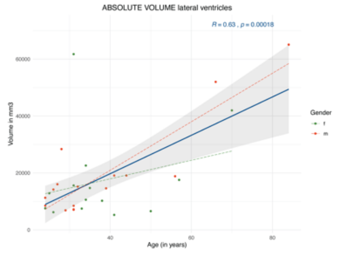


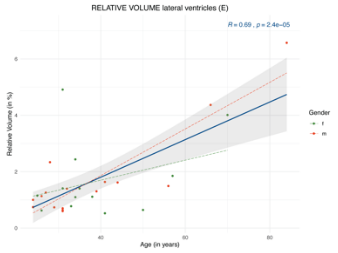


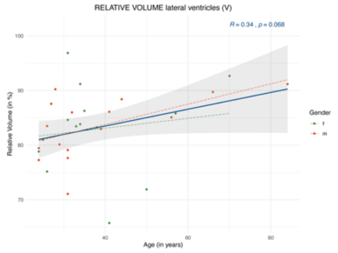


##### FRONTAL HORN

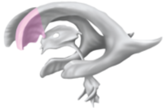

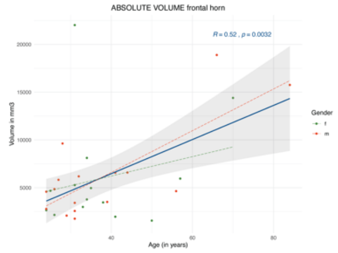


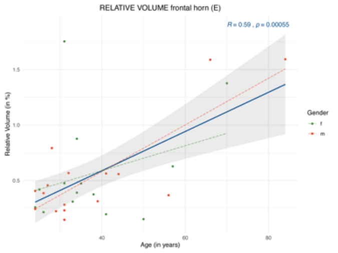


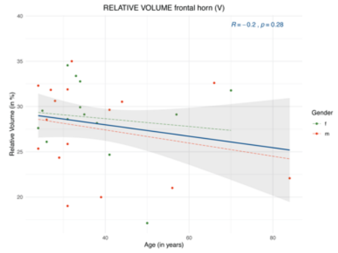


##### BODY

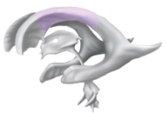

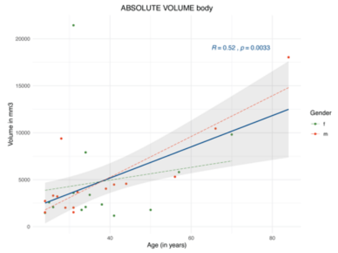


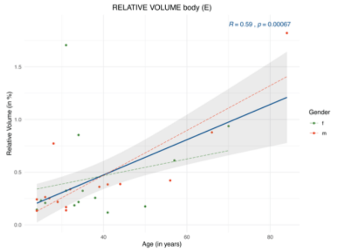


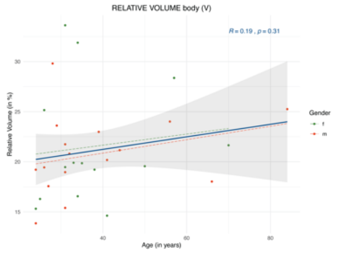

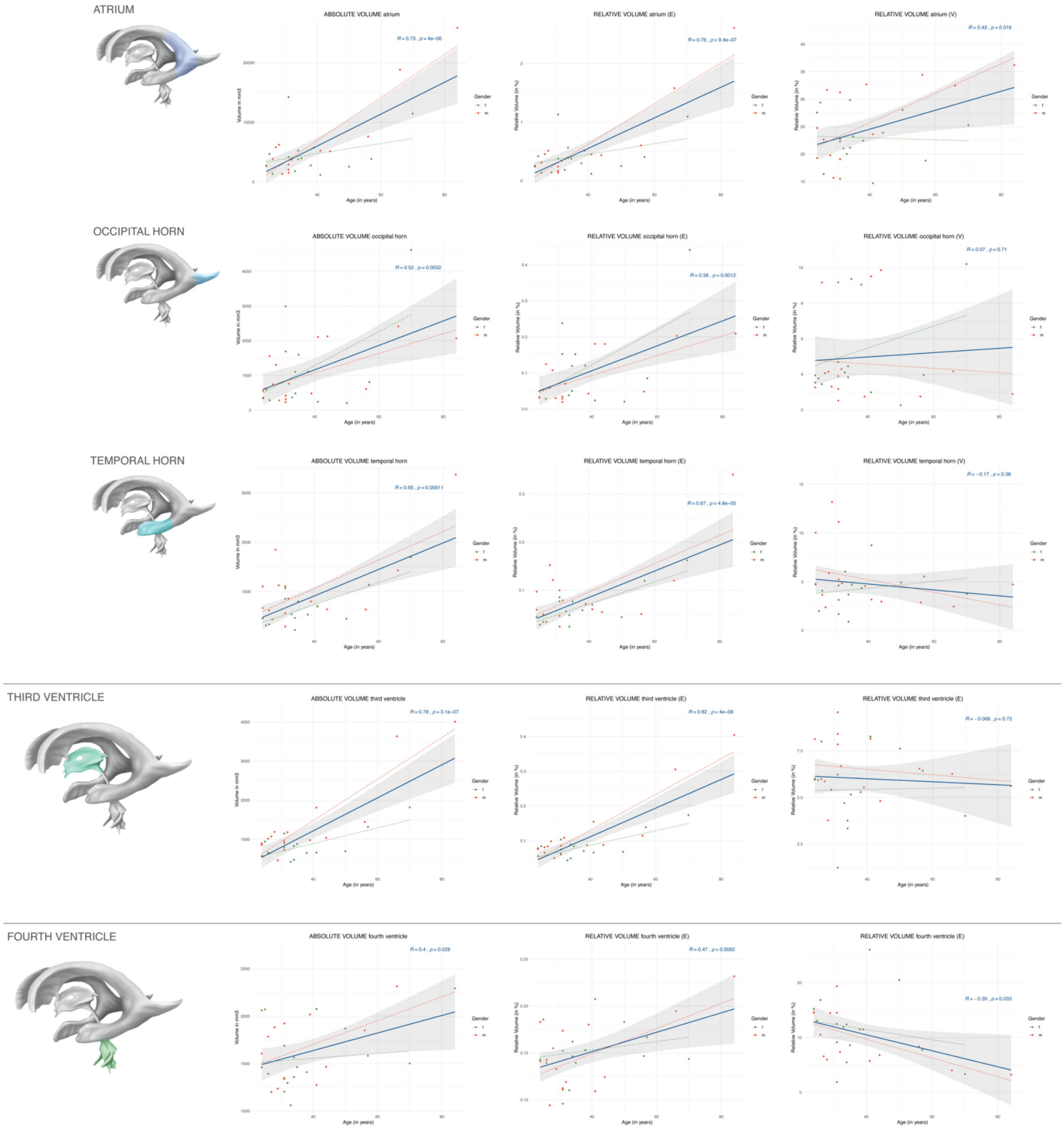


**Supplemental Figure 7. Volumes of the ventricular system correlated to age:** Absolute and relative volumes of the anatomical divisions of the ventricular system in correlation to the age of the subjects. The specific anatomical structures are illustrated schematically. For the total ventricular system, the left diagram displays the correlation between absolute volume and age, the right diagram the correlation between relative volume normalized to the total individual encephalic volume (without ventricles) and age. For each anatomical division of the ventricular system, the left diagram gives the correlation between absolute volume and age, the middle diagram the correlation between relative volume normalized to the total encephalic volume (without ventricles) and age, and the right diagram the correlation between relative volume normalized to the total ventricular volume and age. Color-coding provides a stratification by gender. *Abbreviations*: f = female; m = male.

#### Supplemental Tables

**Supplemental Table 1. Neuroanatomical parcellation algorithm and corresponding segmentation method:** The individual parcellation units are listed with the corresponding anatomical definition and segmentation method. *Abbreviations: A = axial; C = coronal; M = manual; S = sagittal; SA = semi-automated.*

| ANATOMICAL STRUCTURE | ANATOMICAL SPECIFICATION / BOUNDARIES | SEGMENTATION METHOD |
|---|---|---|
| **CEREBRAL LOBES** | *See figure 1A* | |
| **Frontal lobe** | Rostral: Anterior parolfactory sulcus;<br>Caudal: Precentral sulcus (lateral surface), paracentral sulcus (medial surface);<br>Ventral: -<br>Dorsal: -<br>Medial: Cingulate sulcus;<br>Lateral: Anterior periinsular sulcus (anterior), superior periinsular sulcus (posterior). | **SA** |
| **Central lobe** | Rostral: Precentral sulcus (lateral surface), paracentral sulcus (medial surface);<br>Caudal: Postcentral sulcus (lateral surface), marginal sulcus (medial surface);<br>Ventral: -<br>Dorsal: -<br>Medial: Cingulate sulcus;<br>Lateral: Superior periinsular sulcus. | **SA** |
| **Parietal lobe** | Rostral: Postcentral sulcus (lateral surface), marginal sulcus (medial surface);<br>Caudal: Parieto-occipital sulcus (medial surface), parieto-occipital line (lateral surface: line following the anterior occipital sulcus connecting the superior end of the parieto-occipital sulcus and the preoccipital notch);<br>Ventral: -<br>Dorsal: -<br>Medial: Subparietal sulcus<br>Lateral: Parieto-temporal line (arbitrary line between the posterior ramus of the Sylvian fissure and the preoccipital notch). | **SA** |
| **Occipital lobe** | Rostral: Parieto-occipital sulcus (medial surface), parieto-occipital line (lateral surface: line following the anterior occipital sulcus connecting the superior end of the parieto-occipital sulcus and the preoccipital notch); Occipitotemporal line (inferior surface: arbitrary line connecting the inferior end of the parieto-occipital sulcus and the preoccipital notch);<br>Caudal: -<br>Ventral: -<br>Dorsal: -<br>Medial: -<br>Lateral: - | **SA** |

| | | |
|---|---|---|
| **Temporal lobe** | Rostral: -<br><br>Caudal: Parieto-temporal line (lateral surface: arbitrary line between the posterior ramus of the Sylvian fissure and the preoccipital notch), occipitotemporal line (inferior surface: arbitrary line connecting the inferior end of the parieto-occipital sulcus and the preoccipital notch);<br><br>Ventral: -<br><br>Dorsal: Inferior periinsular sulcus;<br><br>Medial: Collateral sulcus (posterior), rhinal sulcus (anterior);<br><br>Lateral: - | **SA** |
| **Insular lobe** | Rostral: Anterior periinsular sulcus;<br><br>Caudal: Posterior insular point;<br><br>Ventral: Posterior periinsular sulcus;<br><br>Dorsal: Superior periinsular sulcus;<br><br>Medial: Limen insulae;<br><br>Lateral: - | **SA** |
| **Limbic lobe** | Rostral: Anterior parolfactory sulcus (anterior), lamina terminalis (posterior);<br><br>Caudal: Subparietal sulcus;<br><br>Ventral: Collateral sulcus (posterior), rhinal sulcus (anterior);<br><br>Dorsal: Cingulate sulcus;<br><br>Medial: Pericallosal sulcus;<br><br>Lateral: Limen insulae. | **SA** |
| **CEREBRAL GYRAL SEGMENTS** | *See figure 2A* | |
| **Frontal pole** | Anterior most aspect of the medial, lateral and inferior surface of the frontal lobe containing the superior, middle and inferior frontopolar gyri;<br><br>Caudal: Superior frontal gyrus, medial frontal gyrus, rostral gyrus, gyrus rectus and anterior orbital gyrus. | **SA** |
| **Superior frontal gyrus** | Rostral: Frontal pole, rostral gyrus;<br><br>Caudal: Precentral sulcus (lateral surface), paracentral sulcus (medial surface);<br><br>Ventral: -<br><br>Dorsal: -<br><br>Medial: Cingulate sulcus;<br><br>Lateral: Superior frontal sulcus. | **SA** |
| **Middle frontal gyrus** | Rostral: Frontal pole;<br><br>Caudal: Precentral sulcus;<br><br>Ventral: -<br><br>Dorsal: -<br><br>Medial: Superior frontal sulcus;<br><br>Lateral: Inferior frontal sulcus. | **SA** |

| | | |
|---|---|---|
| **Inferior frontal gyrus, orbital part** | Rostral: Frontoorbital sulcus;<br>Caudal: Horizonal ramus of the Sylvian fissure;<br>Ventral: Anterior periinsular sulcus;<br>Dorsal: Inferior frontal sulcus;<br>Medial: -<br>Lateral: - | **SA** |
| **Inferior frontal gyrus, triangular part** | Rostral: Horizonal ramus of the Sylvian fissure;<br>Caudal: Ascending ramus of the Sylvian fissure;<br>Ventral: Anterior insular point;<br>Dorsal: Inferior frontal sulcus;<br>Medial: -<br>Lateral: - | **SA** |
| **Inferior frontal gyrus, opercular part** | Rostral: Ascending ramus of the Sylvian fissure;<br>Caudal: Precentral sulcus;<br>Ventral: Superior periinsular sulcus;<br>Dorsal: Inferior frontal sulcus;<br>Medial: -<br>Lateral: - | **SA** |
| **Anterior orbital gyrus** | Rostral: Frontal pole;<br>Caudal: Transverse orbital sulcus;<br>Ventral: -<br>Dorsal: -<br>Medial: Medial orbital sulcus (rostral part)<br>Lateral: Lateral orbital sulcus (caudal part) | **M** |
| **Medial orbital gyrus** | Rostral: Frontal pole;<br>Caudal: Transverse insular gyrus, anterior periinsular sulcus;<br>Ventral: -<br>Dorsal: -<br>Medial: Olfactory sulcus;<br>Lateral: Medial orbital sulcus. | **M** |
| **Lateral orbital gyrus** | Rostral: Frontal pole;<br>Caudal: Anterior periinsular sulcus;<br>Ventral: -<br>Dorsal: -<br>Medial: Lateral orbital sulcus;<br>Lateral: Frontoorbital sulcus. | **M** |

| | | |
|---|---|---|
| **Posterior orbital gyrus** | Rostral: Transverse orbital sulcus; | **M** |
| | Caudal: Transverse insular gyrus, anterior periinsular sulcus; | |
| | Ventral: - | |
| | Dorsal: - | |
| | Medial: Olfactory sulcus; | |
| | Lateral: Frontoorbital sulcus. | |
| **Gyrus rectus** | Rostral: Frontal pole; | **M** |
| | Caudal: Anterior parolfactory sulcus; | |
| | Ventral: - | |
| | Dorsal: Inferior rostral sulcus; | |
| | Medial: - | |
| | Lateral: Medial orbital sulcus. | |
| **Rostral gyrus** | Rostral: Frontal pole; | **M** |
| | Caudal: Anterior parolfactory sulcus; | |
| | Ventral: Inferior rostral sulcus; | |
| | Dorsal: Cingulate sulcus; | |
| | Medial: - | |
| | Lateral: - | |
| **Subcallosal area** | *Comprising the parolfactory gyrus/gyri and the paraterminal gyrus;* | **M** |
| | Rostral: Anterior parolfactory sulcus; | |
| | Caudal: Lamina terminalis; | |
| | Ventral: Diagonal band of Broca; | |
| | Dorsal: Rostrum of corpus callosum (pericallosal sulcus); | |
| | Medial: - | |
| | Lateral: - | |
| **Precentral gyrus** | Rostral: Precentral sulcus; | **SA** |
| | Caudal: Central (Rolandic) sulcus; | |
| | Ventral: - | |
| | Dorsal: - | |
| | Medial: Interhemispheric fissure; | |
| | Lateral: Subcentral gyrus. | |
| **Postcentral gyrus** | Rostral: Central (Rolandic) sulcus; | **SA** |
| | Caudal: Postcentral sulcus; | |
| | Ventral: - | |
| | Dorsal: - | |
| | Medial: Interhemispheric fissure; | |

| | | |
|---|---|---|
| | Lateral: Subcentral gyrus. | |
| **Paracentral lobule** | Rostral: Paracentral sulcus; | **SA** |
| | Caudal: Marginal sulcus; | |
| | Ventral: Cingulate sulcus; | |
| | Dorsal: Interhemispheric fissure; | |
| | Medial: - | |
| | Lateral: - | |
| **Subcentral gyrus** | Rostral: Anterior subcentral sulcus; | **SA** |
| | Caudal: Posterior subcentral sulcus; | |
| | Ventral: - | |
| | Dorsal: Precentral gyrus (anterior), postcentral sulcus (posterior); | |
| | Medial: Superior periinsular sulcus; | |
| | Lateral: - | |
| **Superior parietal lobule** | Rostral: Postcentral sulcus; | **SA** |
| | Caudal: Parieto-occipital line (line following the anterior occipital sulcus connecting the superior end of the parieto-occipital sulcus and the preoccipital notch); | |
| | Ventral: - | |
| | Dorsal: - | |
| | Medial: Interhemispheric fissure; | |
| | Lateral: Intraparietal sulcus. | |
| **Supramarginal gyrus** | Rostral: Postcentral sulcus (superior), posterior subcentral sulcus (inferior); | **SA** |
| | Caudal: Intermediate sulcus of Jensen; | |
| | Ventral: Superior periinsular sulcus, posterior insular point, inferior periinsular sulcus; | |
| | Dorsal: - | |
| | Medial: Intraparietal sulcus; | |
| | Lateral: Parieto-temporal line (arbitrary line between the posterior ramus of the Sylvian fissure and the preoccipital notch). | |
| **Angular gyrus** | Rostral: Intermediate sulcus of Jensen; | **SA** |
| | Caudal: Parieto-occipital line (line following the anterior occipital sulcus connecting the superior end of the parieto-occipital sulcus and the preoccipital notch); | |
| | Ventral: - | |
| | Dorsal: - | |
| | Medial: Intraparietal sulcus; | |
| | Lateral: Parieto-temporal line (arbitrary line between the posterior ramus of the Sylvian fissure and the preoccipital notch). | |
| **Precuneus** | Rostral: Marginal sulcus; | **SA** |
| | Caudal: Parieto-occipital sulcus; | |
| | Ventral: Subparietal sulcus; | |

| | | |
|---|---|---|
| | Dorsal: Interhemispheric fissure;<br>Medial: -<br>Lateral: - | |
| **Cuneus** | Rostral: Parieto-occipital sulcus;<br>Caudal: Interhemispheric fissure;<br>Ventral: Calcarine sulcus;<br>Dorsal: Interhemispheric fissure;<br>Medial: -<br>Lateral: Interhemispheric fissure. | **SA** |
| **Superior occipital gyrus** | Rostral: Parieto-occipital line (line following the anterior occipital sulcus connecting the superior end of the parieto-occipital sulcus and the preoccipital notch);<br>Caudal: Occipital pole;<br>Ventral: -<br>Dorsal: -<br>Medial: Interhemispheric fissure;<br>Lateral: Intraoccipital sulcus. | **SA** |
| **Middle occipital gyrus** | Rostral: Parieto-occipital line (line following the anterior occipital sulcus connecting the superior end of the parieto-occipital sulcus and the preoccipital notch);<br>Caudal: Occipital pole;<br>Ventral: -<br>Dorsal: -<br>Medial: Intraoccipital sulcus;<br>Lateral: Inferior occipital sulcus. | **SA** |
| **Inferior occipital gyrus** | Rostral: Parieto-occipital line (line following the anterior occipital sulcus connecting the superior end of the parieto-occipital sulcus and the preoccipital notch);<br>Caudal: Occipital pole;<br>Ventral: Retrolingual sulcus;<br>Dorsal: -<br>Medial: Inferior occipital sulcus;<br>Lateral: - | **SA** |
| **Occipital pole** | Posterior most aspect of the medial, lateral and inferior surface of the occipital lobe;<br>Rostral: Superior occipital gyrus, medial occipital gyrus, posterior occipital gyrus, cuneus and lingual gyrus. | **SA** |
| **Lingual gyrus** | Rostral: Truncus fissurae parietooccipitalis et calcarinae;<br>Caudal: Occipital pole;<br>Ventral: -<br>Dorsal: Calcarine sulcus;<br>Medial: - | **SA** |

| | | |
|---|---|---|
| | Lateral: Collateral sulcus. | |
| **Fusiform gyrus** | Rostral: Temporal pole;<br><br>Caudal: Junction between the collateral sulcus (medially) and the (lateral) occipitotemporal sulcus (laterally);<br><br>Ventral: -<br><br>Dorsal: -<br><br>Medial: Collaterals sulcus;<br><br>Lateral: (Lateral) occipitotemporal sulcus. | **SA** |
| **Superior temporal gyrus** | Rostral: Temporal pole;<br><br>Caudal: Parieto-temporal line (arbitrary line between the posterior ramus of the Sylvian fissure and the preoccipital notch);<br><br>Ventral: Superior temporal sulcus;<br><br>Dorsal: Sylvian fissure/Transverse temporal gyri/sulci (of Heschl and Schwalbe);<br><br>Medial: -<br><br>Lateral: - | **SA** |
| **Middle temporal gyrus** | Rostral: Temporal pole;<br><br>Caudal: Parieto-temporal line (arbitrary line between the posterior ramus of the Sylvian fissure and the preoccipital notch);<br><br>Ventral: Inferior temporal sulcus;<br><br>Dorsal: Superior temporal sulcus;<br><br>Medial: -<br><br>Lateral: - | **SA** |
| **Inferior temporal gyrus** | Rostral: Temporal pole;<br><br>Caudal: Parieto-temporal line (arbitrary line between the posterior ramus of the Sylvian fissure and the preoccipital notch);<br><br>Ventral: -<br><br>Dorsal: Inferior temporal sulcus<br><br>Medial: (Lateral) occipitotemporal sulcus;<br><br>Lateral: - | **SA** |
| **Planum temporale** | *Comprising the posterior transverse temporal gyri of Heschl;*<br><br>Rostral: Transverse temporal gyri/sulci of Schwalbe;<br><br>Caudal: Terminal (ascending and descending) limbs of the Sylvian fissure;<br><br>Ventral: -<br><br>Dorsal: Posterior insular point;<br><br>Medial: Inferior periinsular sulcus;<br><br>Lateral: Sylvian fissure/superior temporal gyrus. | **SA** |
| **Planum polare** | *Comprising the anterior transverse temporal gyri of Schwalbe;*<br><br>Rostral: Temporal pole; | **SA** |

| | | |
|---|---|---|
| | Caudal: Transverse temporal gyri/sulci of Heschl;<br><br>Ventral: -<br><br>Dorsal: Inferior periinsular sulcus;<br><br>Medial: Limen insulae;<br><br>Lateral: Sylvian fissure/superior temporal gyrus. | |
| **Temporal pole** | Anterior most aspect of the medial, lateral, superior and inferior surface of the temporal lobe; Caudal: Superior temporal gyrus, middle temporal gyrus, inferior temporal gyrus, fusiform gyrus, parahippocampal gyurs and planum polare. | **SA** |
| **Short insular gyri** | Rostral: Anterior periinsular sulcus;<br><br>Caudal: Central insular sulcus;<br><br>Ventral: -<br><br>Dorsal: Superior periinsular sulcus;<br><br>Medial: Limen insulae;<br><br>Lateral: Anterior insular point. | **SA** |
| **Long insular gyri** | Rostral: Central insular sulcus;<br><br>Caudal: Posterior insular point;<br><br>Ventral: -<br><br>Dorsal: Superior periinsular sulcus;<br><br>Medial: Limen insulae;<br><br>Lateral: Inferior periinsular sulcus. | **SA** |
| **Parahippocampal gyrus** | Rostral: Rhinal sulcus, temporal incisure;<br><br>Caudal: Truncus fissurae parietooccipitalis et calcarinae;<br><br>Ventral: -<br><br>Dorsal: Parasubiculum;<br><br>Medial: -<br><br>Lateral: Collateral sulcus, rhinal sulcus. | **SA** |
| **Cingulate gyrus** | *Divided into an anterior (= ascending), middle (=horizontal) and posterior (= descending) part.*<br><br>Rostral: Anterior parolfactory sulcus;<br><br>Caudal: Truncus fissurae parietooccipitalis et calcarinae;<br><br>Ventral: Pericallosal sulcus;<br><br>Dorsal: Cingulate sulcus, subparietal sulcus;<br><br>Medial: -<br><br>Lateral: - | **SA** |
| **CENTRAL PROSENCEPHALON** | *See figure 3A* | |
| **Corpus callosum** | *Composed of the rostrum, genu, trunk/body and splenium.*<br><br>Rostral: Lamina terminalis; | **SA** |

| | | |
|---|---|---|
| | Caudal: - | |
| | Ventral: Lateral ventricle; | |
| | Dorsal: Pericallosal sulcus; | |
| | Medial: - | |
| | Lateral: - | |
| **Claustrum** | Rostral: Rostral external – extreme capsule junction; | **M** |
| | Caudal: Caudal external – extreme capsule junction; | |
| | Ventral: Ventral external – extreme capsule junction; | |
| | Dorsal: Dorsal external – extreme capsule junction; | |
| | Medial: External capsule; | |
| | Lateral: Extreme capsule. | |
| **Putamen** | Rostral: Internal capsule; | **SA** |
| | Caudal: Internal capsule; | |
| | Ventral: Internal capsule; | |
| | Dorsal: Internal capsule; | |
| | Medial: Globus pallidus; | |
| | Lateral: External capsule. | |
| **Caudate nucleus** | *Composed of a head, body and tail.* | **SA** |
| | Bordered by the lateral ventricle and the internal capsule. | |
| **Globus pallidus** | *Composed of an internal and external part.* | **SA** |
| | Rostral: Internal capsule; | |
| | Caudal: Internal caplsue; | |
| | Ventral: Internal capsule; | |
| | Dorsal: Internal capsule; | |
| | Medial: Internal capsule; | |
| | Lateral: Putamen. | |
| **Internal capsule** | Rostral: Caudate nucleus | **M** |
| | Caudal: Thalamus; | |
| | Ventral: Mesencephalon; | |
| | Dorsal: Caudate nucleus; | |
| | Medial: Caudate nucleus (anterior), thalamus (posterior), | |
| | Lateral: Globus pallidus, putamen. | |
| **Innominate Substance** | Rostral: Olfactory striae; | **M** |
| | Caudal: Optic tract (postero-medially), endorhinal sulcus (postero-laterally); | |
| | Ventral: - | |
| | Dorsal: Anterior commissure, globus pallidus, putamen; | |

| | | |
|---|---|---|
| | Medial: Interhemispheric fissue;<br><br>Lateral: Limen insulae. | |
| **Hypothalamus** | Rostral: Lamina terminalis;<br><br>Caudal: Mesencephalon;<br><br>Ventral: Floor of third ventricle;<br><br>Dorsal: Hypothalamic sulcus;<br><br>Medial: Third ventricle;<br><br>Lateral: Innominate substance, anterior commissure, inferior thalamic peduncle. | **SA** |
| **Thalamus** | Rostral: Foramen of Monro;<br><br>Caudal: Velum interpositum cistern;<br><br>Ventral: Mesencephalon;<br><br>Dorsal: Lateral ventricle;<br><br>Medial: Third ventricle;<br><br>Lateral: Internal capsule. | **SA** |
| **Hippocampus** | *Comprising the parasubiculum, presubiculum, subiculum, dentate gyrus, CA1-CA3 field of Ammon's horn.*<br><br>Rostral: Amygdala;<br><br>Caudal: Fornix;<br><br>Ventral: Hippocampal sulcus;<br><br>Dorsal: temporal horn of lateral ventricle, choroid fissure;<br><br>Medial: Crural cistern:<br><br>Lateral: Temporal horn of lateral ventricle. | **SA** |
| **Amygdala** | Rostral: Limen insulae;<br><br>Caudal: Temporal horn of lateral ventricle, hippocampus;<br><br>Ventral: Temporal horn of lateral ventricle, hippocampus;<br><br>Dorsal: Crural cistern, innominate substance;<br><br>Medial: Crural cistern, innominate substance;<br><br>Lateral: Limen insulae, temporal horn of lateral ventricle. | **SA** |
| **BRAINSTEM** | *See figure 4A* | **SA** |
| **Mesencephalon** | Rostral: Mamillary bodies; optic tract, lateral geniculate body, pulvinar thalami, posterior commissure;<br><br>Caudal: Pontomesencephalic sulcus. | **M** |
| **Pons** | Rostral: Pontomesencephalic sulcus;<br><br>Caudal: Pontomedullary sulcus. | **M** |
| **Medulla oblongata** | Rostral: Pontomedullary sulcus;<br><br>Caudal: Lower border of pyramidal decussation. | **M** |
| **CEREBELLUM** | *See figure 4A* | **SA** |

| | | |
|---|---|---|
| **Cerebellar lobes** | | |
| **Anterior** | Comprising the vinculum and lingula (not segmented in this study due to the small size and indistinct identifiability on the MRI), central lobule and ala lobuli centralis, culmen and anterior quadrangular lobule *(definitions below)*. Separated from the medial cerebellar lobe by the primary cerebellar fissure. | **M** |
| **Medial** | Comprising the declive and posterior quadrangular lobule, folium and superior semilunar lobule *(definitions below)*. Separated from the posterior cerebellar lobe by the horizontal cerebellar fissure . | **M** |
| **Posterior** | Comprising the tuber and inferior semilunar/gracile lobule; pyramis and biventer lobule, uvula and tonsil *(definitions below)*. Separated from the flocculonodular cerebellar lobe by the posterolateral cerebellar fissure. | **M** |
| **Flocculonodular** | Comprising the nodulus and flocculus *(definitions below)*. | **M** |
| **Vermian lobules** | Bilateral: Paravermal fissure. | |
| **Central** | Rostral: Precentral cerebellar fissure; | **M** |
| | Caudal: Preculminate cerebellar fissure. | |
| **Culmen** | Rostral: Preculminate cerebellar fissure; | **M** |
| | Caudal: Primary cerebellar fissure. | |
| **Declive** | Rostral: Primary cerebellar fissure; | **M** |
| | Caudal: Postclival (posterior superior) cerebellar fissure. | |
| **Folium** | Rostral: Postclival (posterior superior) cerebellar fissure; | **M** |
| | Caudal: Horizontal cerebellar fissure. | |
| **Tuber** | Rostral: Horizontal cerebellar fissure; | **M** |
| | Caudal: Prepyramidal (prebiventer) cerebellar fissure. | |
| **Pyramid** | Rostral: Prepyramidal (prebiventer) cerebellar fissure; | **M** |
| | Caudal: Secondary (postpyramidal) cerebellar fissure. | |
| **Uvula** | Rostral: Secondary (postpyramidal) cerebellar fissure; | **M** |
| | Caudal: Posterolateral fissure. | |
| **Nodule** | Rostral: Posterolateral fissure; | **M** |
| | Caudal: Inferior medullary velum. | |
| **Cerebellar hemispheric lobules** | Bimedial: Paravermal fissure | |
| **Ala lobuli centralis** | Rostral: Precentral cerebellar fissure; | **M** |
| | Caudal: Preculminate cerebellar fissure. | |
| **Anterior quadrangular lobule** | Rostral: Preculminate cerebellar fissure; | **M** |
| | Caudal: Primary cerebellar fissure. | |
| **Posterior quadrangular lobule** | Rostral: Primary cerebellar fissure; | **M** |
| | Caudal: Postclival (posterior superior) cerebellar fissure. | |
| **Superior semilunar lobule** | Rostral: Postclival (posterior superior) cerebellar fissure; | **M** |
| | Caudal: Horizontal cerebellar fissure. | |
| **Inferior semilunar / gracile lobule** | Rostral: Horizontal cerebellar fissure; | **M** |

| | | |
|---|---|---|
| | Caudal: Prepyramidal (prebiventer) cerebellar fissure. | |
| **Biventer lobule** | Rostral: Prepyramidal (prebiventer) cerebellar fissure; | **M** |
| | Caudal: Secondary (postpyramidal) cerebellar fissure. | |
| **Tonsilla** | Rostral: Secondary (postpyramidal) cerebellar fissure; | **M** |
| | Caudal: Posterolateral fissure. | |
| **Flocculus** | Rostral: Posterolateral fissure; | **M** |
| | Caudal: Inferior medullary velum. | |
| **VENTRICULAR SYSTEM** | *See figure 5A* | **SA** |
| **Lateral ventricles** | Ventricular system proximal to the foramina of Monroe. | **SA** |
| **Frontal horn** | Caudal: Coronal plane through the foramina of Monroe; | **M** |
| | Ventral: Foramina of Monroe. | |
| **Body** | Rostral: Coronal plane through the foramina of Monroe; | **M** |
| | Caudal: Coronal plane through the caudal pole of the thalamus. | |
| **Atrium** | Rostral/ventral: Coronal plane through the caudal pole of the thalamus; | **M** |
| | Caudal: Coronal plane through the rostral tip of the calcar avis; | |
| **Occipital horn** | Rostral: Coronal plane through the rostral tip of the calcar avis. | **M** |
| **Temporal horn** | Caudal: Coronal plane through the caudal pole of the thalamus. | **M** |
| **Third ventricle** | Ventricular system distal to the foramina of Monro and proximal to the cerebral aqueduct. | **SA** |
| **Fourth ventricle** | Ventricular system distal to the cerebral aqueduct and proximal to the foramina of Magendie/Luschkae. | **SA** |
| **Apex** | Caudal: Axial plane through the rostral tip of the limiting sulcus. | **M** |
| **Lateral recess** | Medial: Sagittal plane through the limiting sulcus. | **M** |
| **Obex** | Rostral: Axial plane through the caudal tip of the limiting sulcus. | **M** |
| **Fastigium** | Basal: Coronal plane through the rostral tip of the cerebellar nodule. | **M** |

**Supplemental Table 2. Volumes of general anatomical structures:** The absolute and relative volumes of the cerebral lobes, the basal ganglia, the diencephalon as well as the brainstem and cerebellum are given. Absolute volumes are provided in mm$^3$, relative volumes in % normalized to the total individual encephalic volume (without ventricles). The volumes are given as mean and standard deviation (SD). The relative standard deviation (RSD) corresponds to the coefficient of variation and results from the proportion of the SD to the mean (in %). A stratification by gender is given for absolute and relative volumes. The provided p-value reflects the level of evidence for a gender-difference based on two-sample t-test statistics and needs to be interpreted in consideration of multiple testing.

| **Anatomical Structure** | **Absolute Volumes in mm$^3$** [mean (SD, RSD)] | | | | **Relative Volumes in %** [mean (SD, RSD)] | | | |
|---|---|---|---|---|---|---|---|---|
| | **Total**, n = 30 | **Female**, n = 14 | **Male**, n = 16 | ***p*** | **Total**, n = 30 | **Female**, n = 14 | **Male**, n = 16 | ***p*** |
| **Frontal lobe** | 199’480 (24’676, 12.4%) | 186’347 (19’262, 10.3%) | 210’971 (23’534, 11.2%) | *0.004* | **18.23** (0.96, 5.3%) | 18.17 (0.74, 4.1%) | 18.27 (1.14, 6.2%) | *0.784* |
| **Central lobe** | 81’517 (9’208, 11.3%) | 76’746 (8’368, 10.9%) | 85’692 (7’962, 9.3%) | *0.006* | **7.47** (0.52, 7.0%) | 7.49 (0.43, 5.7%) | 7.45 (0.61, 8.2%) | *0.843* |
| **Parietal lobe** | 129’216 (16’770, 13%) | 120’829 (15’806, 13.1%) | 136’554 (14’287, 10.5%) | *0.008* | **11.82** (0.89, 7.5%) | 11.77 (0.78, 6.7%) | 11.86 (1.00, 8.4%) | *0.784* |
| **Occipital lobe** | 75’911 (8’885, 11.7%) | 70’930 (5’286, 7.5%) | 80’268 (9’219, 11.5%) | *0.002* | **6.95** (0.47, 6.7%) | 6.94 (0.49, 7.0%) | 6.95 (0.46, 6.7%) | *0.956* |
| **Temporal lobe** | 116’477 (13’868, 11.9%) | 109’342 (9’774, 8.9%) | 122’720 (14’140, 11.5%) | *0.006* | **10.66** (0.71, 6.6%) | 10.67 (0.32, 3.0%) | 10.65 (0.94, 8.8%) | *0.929* |
| **Insular lobe** | 24’547 (3’015, 12.3%) | 22’746 (2’321, 10.2%) | 26’123 (2’688, 10.3%) | *0.001* | **2.25** (0.18, 8.2%) | 2.22 (0.17, 7.7%) | 2.27 (0.20, 8.7%) | *0.501* |
| **Limbic lobe** | 75’816 (9’372, 12.4%) | 71’385 (8’463, 11.9%) | 79’694 (8’567, 10.7%) | *0.013* | **6.93** (0.38, 5.5%) | 6.95 (0.36, 5.1%) | 6.90 (0.41, 6.0%) | *0.720* |
| **Basal ganglia** | 22’117 (2’979, 13.5%) | 21’023 (2’276, 10.8%) | 23’075 (3’251, 14.1%) | *0.058* | **2.03** (0.23, 11.4%) | 2.05 (0.18, 8.9%) | 2.01 (0.27, 13.5%) | *0.573* |
| **Diencephalon** | 22’569 (2’269, 10.1%) | 21’697 (1’747, 8.1%) | 23’331 (2’445, 10.5%) | *0.047* | **2.07** (0.13, 6.4%) | 2.12 (0.11, 5.0%) | 2.02 (0.14, 7.0%) | *0.045* |
| **Brainstem** | 28’072 (3’293, 11.7%) | 26’808 (3’261, 12.2%) | 29’179 (2’991, 10.3%) | *0.047* | **2.57** (0.24, 9.2%) | 2.62 (0.26, 9.9%) | 2.53 (0.22, 8.5%) | *0.335* |
| **Cerebellum** | 116’733 (12’623, 10.8%) | 111’617 (12’961, 11.6%) | 121’210 (10’802, 8.9%) | *0.035* | **10.73** (1.17, 10.9%) | 10.93 (1.22, 11.2%) | 10.56 (1.13, 10.7%) | *0.397* |

**Supplemental Table 3. Volumes of the cerebral gyral segments:** The absolute and relative volumes of the cerebral gyral segments are given. Absolute volumes are provided in $mm^3$, relative volumes in % normalized to the total individual encephalic volume (without ventricles). The volumes are given as mean and standard deviation (SD). The relative standard deviation (RSD) corresponds to the coefficient of variation and results from the proportion of the SD to the mean (in %). A stratification by gender is given for absolute and relative volumes. The provided p-value reflects the level of evidence for a gender-difference based on two-sample t-test statistics and needs to be interpreted in consideration of multiple testing. *Abbreviations*: ANG = angular gyrus; F1 = superior frontal gyrus; F2 = middle frontal gyrus; F3 = inferior frontal gyrus; O1 = superior occipital gyrus; O2 = middle occipital gyrus; O3 = inferior occipital gyrus; PHG = parahippocampal gyrus; SCA = subcallosal area; SMG = supramarginal gyrus; SPL = superior parietal lobule; T1 = superior temporal gyrus; T2 = middle temporal gyrus; T3 = inferior temporal gyrus.

| **Anatomical Structure** | **Absolute Volumes in $mm^3$** [mean (SD, RSD)] | | | | **Relative Volumes in %** [mean (SD, RSD)] | | | |
|---|---|---|---|---|---|---|---|---|
| | **Total**, n = 30 | **Female**, n = 14 | **Male**, n = 16 | ***p*** | **Total**, n = 30 | **Female**, n = 14 | **Male**, n = 16 | ***p*** |
| **Frontal pole** | 4’711 (976, 20.7%) | 4’369 (706, 16.2%) | 5’009 (1’099, 21.9%) | *0.072* | **0.43** (0.074, 17.1%) | 0.43 (0.071, 16.7%) | 0.43 (0.078, 18.0%) | *0.861* |
| **F1** | 68’776 (9’196, 13.4%) | 64’718 (8’455, 13.1%) | 72’328 (8’527, 11.8%) | *0.021* | **6.28** (0.39, 6.2%) | 6.30 (0.36, 5.8%) | 6.26 (0.42, 6.6%) | *0.788* |
| **F2** | 58’458 (7’690, 13.2%) | 55’367 (7’298, 13.2%) | 61’164 (7’172, 11.7%) | *0.037* | **5.34** (0.32, 6.0%) | 5.39 (0.31, 5.7%) | 5.29 (0.34, 6.3%) | *0.428* |
| **F3 orbital** | 3’917 (614, 15.7%) | 3’666 (565, 15.4%) | 4’136 (584, 14.1%) | *0.034* | **0.36** (0.040, 11.2%) | 0.36 (0.039, 10.8%) | 0.36 (0.043, 11.9%) | *0.913* |
| **F3 triangular** | 11’689 (1’899, 16.2%) | 10’846 (1’039, 9.6%) | 12’427 (2’189, 17.6%) | *0.020* | **1.07** (0.15, 13.6%) | 1.06 (0.12, 11.3%) | 1.08 (0.17, 15.6%) | *0.793* |
| **F3 opercular** | 13’949 (2’022, 14.5%) | 12’752 (1’540, 12.1%) | 14’996 (1’828, 12.2%) | *0.001* | **1.28** (0.15, 11.7%) | 1.25 (0.16, 12.6%) | 1.30 (0.14, 11.0%) | *0.339* |
| **Anterior orbital** | 3’349 (604, 18.0%) | 3’087 (245, 7.9%) | 3’579 (730, 20.4%) | *0.023* | **0.31** (0.037, 12.2%) | 0.30 (0.016, 5.5%) | 0.31 (0.049, 16.0%) | *0.586* |
| **Medial orbital** | 7’566 (1’369, 18.1%) | 6’973 (559, 8.0%) | 8’085 (1’655, 20.5%) | *0.024* | **0.69** (0.085, 12.3%) | 0.68 (0.037, 5.4%) | 0.70 (0.11, 16.0%) | *0.586* |
| **Lateral orbital** | 6’064 (1’102, 18.2%) | 5’591 (456, 8.2%) | 6’477 (1’333, 20.6%) | *0.025* | **0.55** (0.068, 12.4%) | 0.55 (0.030, 5.6%) | 0.56 (0.090, 16.1%) | *0.600* |
| **Posterior orbital** | 7’421 (1’341, 18.1%) | 6’845 (546, 8.0%) | 7’924 (1’625, 20.5%) | *0.025* | **0.68** (0.083, 12.3%) | 0.67 (0.037, 5.5%) | 0.68 (0.11, 16.1%) | *0.612* |
| **Rectus** | 9’987 (4’584, 45.9%) | 8’721 (747, 8.6%) | 11’094 (6’111, 55.1%) | *0.161* | **0.91** (0.40, 43.9%) | 0.85 (0.066, 7.8%) | 0.97 (0.55, 56.8%) | *0.452* |
| **Rostral** | 3’593 (380, 10.6%) | 3’413 (296, 8.7%) | 3’751 (383, 10.2%) | *0.012* | **0.33** (0.025, 7.6%) | 0.33 (0.026, 7.9%) | 0.33 (0.024, 7.3%) | *0.357* |
| **Precentral** | 39’114 (4’348, 11.1%) | 36’944 (3’428, 9.3%) | 41’013 (4’258, 10.4%) | *0.008* | **3.59** (0.30, 8.4%) | 3.61 (0.25, 6.8%) | 3.57 (0.35, 9.8%) | *0.697* |
| **Postcentral** | 22’308 (3’281, 14.7%) | 20’938 (3’435, 16.4%) | 23’507 (2’701, 11.5%) | *0.030* | **2.04** (0.18, 8.8%) | 2.04 (0.19, 9.3%) | 2.04 (0.18, 8.7%) | *0.953* |

| | | | | | | | | |
|---|---|---|---|---|---|---|---|---|
| **Paracentral lobule** | 14’037 (1’770, 12.6%) | 13’362 (1’786, 13.4%) | 14’628 (1’579, 10.8%) | *0.049* | **1.29** (0.14, 10.9%) | 1.30 (0.14, 11.0%) | 1.27 (0.14, 11.0%) | *0.538* |
| **Subcentral** | 6’058 (1’028, 17.0%) | 5’502 (886, 16.1%) | 6’544 (906, 13.8%) | *0.004* | **0.55** (0.070, 12.7%) | 0.54 (0.057, 10.6%) | 0.57 (0.078, 13.8%) | *0.199* |
| **SPL** | 33’315 (4’452, 13.4%) | 31’813 (4’850, 15.2%) | 34’630 (3’741, 10.8%) | *0.084* | **3.05** (0.30, 9.8%) | 3.10 (0.28, 9.0%) | 3.01 (0.32, 10.7%) | *0.463* |
| **SMG** | 30’708 (4’786, 15.6%) | 28’641 (4’366, 15.2%) | 32’516 (4’504, 13.9%) | *0.024* | **2.81** (0.30, 10.6%) | 2.79 (0.26, 9.2%) | 2.82 (0.34, 11.9%) | *0.756* |
| **ANG** | 34’333 (4’761, 13.9%) | 31’865 (4’552, 14.3%) | 36’494 (3’898, 10.7%) | *0.006* | **3.14** (0.30, 9.5%) | 3.11 (0.33, 10.7%) | 3.17 (0.27, 8.5%) | *0.582* |
| **Precuneus** | 30’859 (4’303, 13.9%) | 28’510 (3’789, 13.3%) | 32’915 (3’700, 11.2%) | *0.003* | **2.82** (0.22, 7.8%) | 2.78 (0.21, 7.5%) | 2.85 (0.23, 8.1%) | *0.341* |
| **Cuneus** | 10’063 (1’462, 14.5%) | 9’355 (1’040, 11.1%) | 10’683 (1’522, 14.2%) | *0.010* | **0.92** (0.11, 12.0%) | 0.92 (0.11, 11.9%) | 0.93 (0.12, 12.5%) | *0.803* |
| **O1** | 9’149 (1’312, 14.3%) | 8’299 (926, 11.2%) | 9’894 (1’149, 11.6%) | *<0.001* | **0.84** (0.080, 9.5%) | 0.81 (0.073, 9.0%) | 0.86 (0.081, 9.4%) | *0.100* |
| **O2** | 15’787 (2’400, 15.2%) | 14’684 (1’913, 13.0%) | 16’752 (2’415, 14.4%) | *0.016* | **1.44** (0.14, 9.7%) | 1.43 (0.16, 10.8%) | 1.45 (0.13, 8.9%) | *0.792* |
| **O3** | 9’264 (1’669, 18.0%) | 8’261 (1’177, 14.2%) | 10’141 (1’557, 15.3%) | *0.001* | **0.85** (0.11, 13.4%) | 0.81 (0.12, 14.5%) | 0.88 (0.10, 11.8%) | *0.098* |
| **Occipital pole** | 11’367 (1’512, 13.3%) | 10’667 (1’018, 9.5%) | 11’980 (1’632, 13.6%) | *0.015* | **1.04** (0.10, 9.2%) | 1.05 (0.11, 10.8%) | 1.04 (0.081, 7.9%) | *0.786* |
| **Lingual** | 20’280 (2’734, 13.5%) | 19’665 (2’166, 11.0%) | 20’819 (3’118, 15.0%) | *0.256* | **1.86** (0.23, 12.4%) | 1.93 (0.23, 11.9%) | 1.80 (0.22, 12.4%) | *0.153* |
| **Fusiform** | 22’196 (3’299, 14.9%) | 20’947 (2’858, 13.6%) | 23’289 (3’351, 14.4%) | *0.050* | **2.03** (0.23, 11.3%) | 2.04 (0.21, 10.5%) | 2.02 (0.25, 12.3%) | *0.799* |
| **Temporal pole** | 25’354 (3’578, 14.1%) | 23’479 (2’924, 12.5%) | 26’994 (3’346, 12.4%) | *0.005* | **2.32** (0.20, 8.5%) | 2.29 (0.17, 7.5%) | 2.34 (0.22, 9.4%) | *0.483* |
| **T1** | 27’247 (3’925, 14.4%) | 24’989 (2’642, 10.6%) | 29’224 (3’848, 13.2%) | *0.002* | **2.49** (0.23, 9.3%) | 2.44 (0.16, 6.7%) | 2.53 (0.28, 10.9%) | *0.265* |
| **T2** | 23’693 (3’139, 13.3%) | 22’876 (2’876, 12.6%) | 24’408 (3’274, 13.4%) | *0.187* | **2.17** (0.22, 10.2%) | 2.23 (0.19, 8.4%) | 2.12 (0.24, 11.5%) | *0.171* |
| **T3** | 5’011 (871, 17.4%) | 4’531 (734, 16.2%) | 5’430 (774 14.3%) | *0.003* | **0.46** (0.057, 12.5%) | 0.44 (0.060, 13.5%) | 0.47 (0.053, 11.2%) | *0.179* |
| **Planum temporale** | 3’078 (771, 25.0%) | 2’715 (410, 15.1%) | 3’396 (879, 25.9%) | *0.013* | **0.28** (0.057, 20.3%) | 0.27 (0.037, 14.0%) | 0.29 (0.069, 23.3%) | *0.174* |
| **Planum polare** | 9’898 (1’848 18.7%) | 9’806 (1’646, 16.8%) | 9’979 (2’059, 20.6%) | *0.803* | **0.91** (0.17, 18.8%) | 0.96 (0.17, 17.3%) | 0.87 (0.17, 19.4%) | *0.135* |
| **Short insular** | 15’405 (2’109, 13.7%) | 14’653 (1’336, 9.1%) | 16’064 (2’461, 15.3%) | *0.066* | **1.41** (0.18, 12.5%) | 1.43 (0.12, 8.4%) | 1.40 (0.22, 15.5%) | *0.591* |
| **Long insular** | 9’142 (1’881, 20.6%) | 8’094 (1’222, 15.1%) | 10’059 (1’904, 18.9%) | *0.003* | **0.83** (0.12, 14.4%) | 0.79 (0.089, 11.3%) | 0.87 (0.13, 15.2%) | *0.060* |
| **SCA** | 2’394 (663, 27.7%) | 2’108 (542, 25.7%) | 2’645 (672, 25.4%) | *0.024* | **0.22** (0.047, 21.7%) | 0.20 (0.044, 21.6%) | 0.23 (0.048, 21.2%) | *0.175* |
| **Cingulate anterior** | 14’420 (2’135, 14.8%) | 13’500 (2’194, 16.2%) | 15’224 (1’779, 11.7%) | *0.025* | **1.32** (0.13, 10.2%) | 1.31 (0.14, 11.0%) | 1.32 (0.13, 9.9%) | *0.879* |
| **Cingulate middle** | 17’135 (2’614, 15.3%) | 16’045 (2’246, 14.0%) | 18’090 (2’599, 14.4%) | *0.030* | **1.56** (0.15, 9.5%) | 1.56 (0.13, 8.0%) | 1.57 (0.17, 10.8%) | *0.951* |

| | | | | | | | | |
|---|---|---|---|---|---|---|---|---|
| **Cingulate posterior** | 20’174 (2’958, 14.7%) | 18’886 (2’680, 14.2%) | 21’302 (2’790, 13.1%) | *0.023* | **1.84** (0.15, 7.9%) | 1.84 (0.13, 7.3%) | 1.84 (0.16, 8.6%) | *0.911* |
| **PHG** | 10’443 (1’653, 15.8%) | 10’013 (1’197, 12.0%) | 10’820 (1’927, 17.8%) | *0.187* | **0.96** (0.12, 12.2%) | 0.98 (0.073, 7.5%) | 0.94 (0.15, 15.5%) | *0.392* |

**Supplemental Table 4. Volumes of central prosencephalic structures:** The absolute and relative volumes of the anatomical white and gray matter structures of the central prosencephalon. Absolute volumes are provided in $mm^3$, relative volumes in % normalized to the total individual encephalic volume (without ventricles). The volumes are given as mean and standard deviation (SD). The relative standard deviation (RSD) corresponds to the coefficient of variation and results from the proportion of the SD to the mean (in %). A stratification by gender is given for absolute and relative volumes. The provided p-value reflects the level of evidence for a gender-difference based on two-sample t-test statistics and needs to be interpreted in consideration of multiple testing.

| **Anatomical Structure** | **Absolute Volumes in $mm^3$** [mean (SD, RSD)] | | | | **Relative Volumes in %** [mean (SD, RSD)] | | | |
|---|---|---|---|---|---|---|---|---|
| | **Total**, n = 30 | **Female**, n = 14 | **Male**, n = 16 | ***p*** | **Total**, n = 30 | **Female**, n = 14 | **Male**, n = 16 | ***p*** |
| **Corpus callosum** | 3’312 (552, 16.7%) | 3’319 (580, 17.5%) | 3’306 (546, 16.5%) | *0.949* | **0.30** (0.047, 15.4%) | 0.32 (0.043, 13.2%) | 0.29 (0.045, 15.6%) | *0.033* |
| **Claustrum** | 1’345 (418, 31.1%) | 1’190 (306, 25.7%) | 1’481 (463, 31.3%) | *0.056* | **0.12** (0.036, 29.2%) | 0.12 (0.026, 22.3%) | 0.13 (0.043, 33.1%) | *0.316* |
| **Putamen** | 11’265 (1’662, 14.8%) | 10’794 (1’243, 11.5%) | 11’677 (1’901, 16.3%) | *0.149* | **1.03** (0.14, 13.4%) | 1.06 (0.10, 9.8%) | 1.02 (0.16, 16.2%) | *0.454* |
| **Caudate** | 7’780 (1’318, 16.9%) | 7’368 (932, 12.6%) | 8’140 (1’520, 18.7%) | *0.111* | **0.71** (0.11, 15.2%) | 0.72 (0.075, 10.4%) | 0.71 (0.13, 18.9%) | *0.788* |
| **Globus pallidum** | 3’073 (592, 19.3%) | 2’861 (331, 11.6%) | 3’258 (711, 21.8%) | *0.066* | **0.28** (0.039, 14.0%) | 0.28 (0.030, 10.8%) | 0.28 (0.047, 16.6%) | *0.941* |
| **Internal capsule** | 10’616 (1’549, 14.6%) | 10’153 (1’655, 16.3%) | 11’022 (1’374, 12.5%) | *0.127* | **0.97** (0.12, 12.1%) | 0.99 (0.13, 12.8%) | 0.96 (0.11, 11.5%) | *0.454* |
| **Innominate substance** | 2’688 (323, 12.0%) | 2’639 (363, 13.8%) | 2’731 (288, 10.5%) | *0.448* | **0.25** (0.036, 14.6%) | 0.26 (0.039, 15.1%) | 0.24 (0.031, 13.1%) | *0.119* |
| **Hypothalamus** | 7’963 (917, 11.5%) | 7’507 (671, 8.9%) | 8’361 (935, 11.2%) | *0.008* | **0.73** (0.048, 6.6%) | 0.73 (0.047, 6.4%) | 0.72 (0.050, 6.9%) | *0.620* |
| **Thalamus** | 14’606 (1’463, 10.0%) | 14’190 (1’205, 8.5%) | 14’970 (1’605, 10.7%) | *0.148* | **1.34** (0.10, 7.6%) | 1.39 (0.08, 5.6%) | 1.30 (0.10, 8.1%) | *0.016* |
| **Hippocampus** | 8’128 (953, 11.7%) | 7’824 (712, 9.1%) | 8’394 (1’074, 12.8%) | *0.103* | **0.74** (0.062, 8.4%) | 0.77 (0.064, 8.3%) | 0.73 (0.057, 7.8%) | *0.086* |
| **Amygdala** | 3’122 (470, 15.0%) | 3’009 (469, 15.6%) | 3’220 (462, 14.4%) | *0.227* | **0.29** (0.040, 14.0%) | 0.29 (0.047, 15.8%) | 0.28 (0.033, 11.9%) | *0.293* |

**Supplemental Table 5. Volumes of brainstem and cerebellar structures:** The absolute and relative volumes of the anatomical structures of the brainstem and the cerebellum. Absolute volumes are provided in $mm^3$, relative volumes in % normalized to the total individual encephalic volume (without ventricles). The volumes are given as mean and standard deviation (SD). The relative standard deviation (RSD) corresponds to the coefficient of variation and results from the proportion of the SD to the mean (in %). A stratification by gender is given for absolute and relative volumes. The provided p-value reflects the level of evidence for a gender-difference based on two-sample t-test statistics and needs to be interpreted in consideration of multiple testing.

| **Anatomical Structure** | **Absolute Volumes in $mm^3$** [mean (SD, RSD)] | | | | **Relative Volumes in %** [mean (SD, RSD)] | | | |
|---|---|---|---|---|---|---|---|---|
| | **Total**, n = 30 | **Female**, n = 14 | **Male**, n = 16 | ***p*** | **Total**, n = 30 | **Female**, n = 14 | **Male**, n = 16 | ***p*** |
| **Mesencephalon** | 10’046 (1’461, 14.5%) | 9’764 (1’207, 12.4%) | 10’292 (1’650, 16.0%) | *0.332* | **0.92** (0.11, 12%) | 0.95 (0.10, 10.8%) | 0.89 (0.11, 12.5%) | *0.116* |
| **Pons** | 15’286 (2’216, 14.5%) | 14’389 (2’401, 16.7%) | 16’071 (1’758, 10.9%) | *0.036* | **1.40** (0.17, 12.3%) | 1.40 (0.20, 14.0%) | 1.40 (0.16, 11.1%) | *0.931* |
| **Medulla oblongata** | 2’741 (368, 13.4%) | 2’655 (274, 10.3%) | 2’816 (429, 15.2%) | *0.238* | **0.25** (0.036, 14.2%) | 0.26 (0.033, 12.7%) | 0.25 (0.038, 15.3%) | *0.238* |
| **Cerebellar peduncles** | 6’781 (962, 14.2%) | 6’443 (696, 10.8%) | 7’076 (1’082, 15.3%) | *0.071* | **0.62** (0.072, 11.6%) | 0.63 (0.070, 11.1%) | 0.61 (0.074, 12.1%) | *0.501* |
| **Vermis** | 5’947 (805, 13.5%) | 5’729 (698, 12.2%) | 6’137 (864, 14.1%) | *0.170* | **0.55** (0.070, 12.9%) | 0.56 (0.070, 12.5%) | 0.53 (0.071, 13.3%) | *0.289* |
| **Hemisphere** | 110’786 (12’112, 10.9%) | 105’887 (12’511, 11.8%) | 115’073 (10’293, 8.9%) | *0.036* | **10.19** (1.12, 11.0%) | 10.37 (1.18, 11.3%) | 10.03 (1.08, 10.8%) | *0.416* |
| **Anterior lobe** | 32’233 (4’776, 14.8%) | 30’952 (5’067, 16.4%) | 33’353 (4’356, 13.1%) | *0.174* | **2.97** (0.50, 16.7%) | 3.03 (0.52, 17.0%) | 2.92 (0.49, 16.7%) | *0.521* |
| **Medial lobe** | 28’792 (4’415, 15.3%) | 28’502 (4’695, 16.5%) | 29’045 (4’293, 14.8%) | *0.743* | **2.64** (0.37, 14.0%) | 2.78 (0.39, 14.2%) | 2.52 (0.30, 12.1%) | *0.048* |
| **Posterior lobe** | 54’645 (7’773, 14.2%) | 51’124 (7’945, 15.5%) | 57’726 (6’357, 11.0%) | *0.017* | **5.02** (0.70, 13.8%) | 5.01 (0.77, 15.5%) | 5.03 (0.64, 12.8%) | *0.932* |
| **Flocculonodular lobe** | 1’064 (150, 14.1%) | 1’040 (172, 16.5%) | 1’085 (131, 12.1%) | *0.422* | **0.098** (0.017, 17.7%) | 0.10 (0.018, 17.8%) | 0.095 (0.017, 17.4%) | *0.259* |
| **Central** | 482 (125, 26.0%) | 494 (139, 28.1%) | 471 (115, 24.5%) | *0.627* | **0.044** (0.012, 27.2%) | 0.048 (0.013, 27.5%) | 0.041 (0.010, 24.4%) | *0.091* |
| **Culmen** | 2’184 (360, 16.5%) | 2’090 (300, 14.4%) | 2’265 (396, 17.5%) | *0.189* | **0.20** (0.032, 15.9%) | 0.20 (0.029, 14.0%) | 0.20 (0.035, 17.7%) | *0.541* |
| **Declive** | 1’031 (189, 18.3%) | 1’010 (222, 22.0%) | 1’049 (158, 15.1%) | *0.577* | **0.095** (0.017, 17.7%) | 0.098 (0.020, 19.9%) | 0.091 (0.013, 14.8%) | *0.246* |
| **Folium** | 440 (120, 27.4%) | 430 (104, 24.2%) | 449 (136, 30.3) | *0.673* | **0.041** (0.011, 27.4%) | 0.042 (0.0093, 22.3%) | 0.039 (0.013, 32.1%) | *0.549* |
| **Tuber** | 387 (97.0, 25.1%) | 359 (74.5, 20.8%) | 412 (110, 26.6%) | *0.143* | **0.036** (0.0085, 23.9%) | 0.035 (0.0083, 23.7%) | 0.036 (0.0088, 24.8%) | *0.913* |
| **Pyramid** | 427 (202, 47.3%) | 368 (138, 37.5%) | 478 (237, 49.6%) | *0.140* | **0.039** (0.017, 43.4%) | 0.037 (0.015, 42.1%) | 0.041 (0.018, 44.8%) | *0.499* |
| **Uvula** | 785 (129, 16.4%) | 768 (121, 15.8%) | 799 (138, 17.2%) | *0.527* | **0.072** (0.012, 16.9%) | 0.075 (0.013, 16.9%) | 0.069 (0.011, 16.3%) | *0.179* |
| **Nodule** | 213 (35.1, 16.5%) | 210 (32.7, 15.5%) | 214 (38.1, 17.8%) | *0.768* | **0.020** (0.0037, 18.9) | 0.021 (0.0038, 18.2) | 0.019 (0.0035, 18.6) | *0.135* |

| | | | | | | | | |
|---|---|---|---|---|---|---|---|---|
| **Ala lobuli centralis** | 11’994 (2’880, 24.0%) | 11’411 (3’164, 27.7%) | 12’504 (2’601, 20.8%) | *0.308* | **1.10** (0.27, 24.8%) | 1.11 (0.29, 25.9%) | 1.10 (0.27, 24.6%) | *0.879* |
| **AQL** | 17’574 (3’822, 21.7%) | 16’957 (3’679, 21.7%) | 18’113 (3’981, 22.0%) | *0.418* | **1.62** (0.38, 23.4%) | 1.67 (0.40, 24.2%) | 1.58 (0.36, 23.1%) | *0.533* |
| **PQL** | 13’124 (2’598, 19.8%) | 12’835 (3’259, 25.4%) | 13’378 (1’924, 14.4%) | *0.577* | **1.20** (0.22, 18.2%) | 1.25 (0.27, 21.5%) | 1.16 (0.16, 14.1%) | *0.311* |
| **SSL** | 14’197 (2’514, 17.7%) | 14’227 (2’164, 15.2%) | 14’170 (2’856, 20.2%) | *0.952* | **1.30** (0.23, 17.6%) | 1.40 (0.23, 16.2%) | 1.22 (0.21, 16.8%) | *0.038* |
| **ISL/gracile** | 34’930 (6’198, 17.7%) | 33’389 (6’276, 18.8%) | 36’279 (5’997, 16.5%) | *0.208* | **3.22** (0.63, 19.5%) | 3.27 (0.59, 18.0%) | 3.18 (0.68, 21.3%) | *0.705* |
| **Biventer** | 12’163 (3’787, 31.1%) | 10’530 (3’582, 34.0%) | 13’593 (3’454, 25.4%) | *0.024* | **1.11** (0.31, 28.2%) | 1.03 (0.36, 35.2%) | 1.17 (0.25, 21.6%) | *0.236* |
| **Tonsilla** | 5’953 (1’338, 22.5%) | 5’710 (1’080, 18.9%) | 6’165 (1’532, 24.8%) | *0.361* | **0.55** (0.12, 21.3%) | 0.56 (0.11, 19.9%) | 0.53 (0.12, 23.1%) | *0.577* |
| **Flocculus** | 851 (139, 16.4%) | 829 (159, 19.2%) | 870 (122, 14.0%) | *0.428* | **0.079** (0.016, 19.7%) | 0.081 (0.016, 19.8%) | 0.076 (0.015, 19.7%) | *0.364* |

**Supplemental Table 6. Volumes of the ventricular system:** The absolute and relative volumes of the anatomical divisions of the ventricular system. Absolute volumes are provided in $mm^3$, relative volumes in % normalized to either the total individual encephalic volume (without ventricles) or the total individual ventricular volume. The volumes are given as mean and standard deviation (SD). The relative standard deviation (RSD) corresponds to the coefficient of variation and results from the proportion of the SD to the mean (in %). A stratification by gender is given for absolute and relative volumes. The provided p-value reflects the level of evidence for a gender-difference based on two-sample t-test statistics and needs to be interpreted in consideration of multiple testing. *Abbreviations*: LV = lateral ventricle.

| **Anatomical Structure** | **Absolute Volumes in $mm^3$** [mean (SD, RSD)] | | | | **Relative Volumes in %– normalized to total encephalic volume** [mean (SD, RSD)] | | | | **Relative Volumes in %– normalized to total ventricular volume** [mean (SD, RSD)] | | | |
|---|---|---|---|---|---|---|---|---|---|---|---|---|
| | **Total**, n = 30 | **Female**, n = 14 | **Male**, n = 16 | ***p*** | **Total**, n = 30 | **Female**, n = 14 | **Male**, n = 16 | ***p*** | **Total**, n = 30 | **Female**, n = 14 | **Male**, n = 16 | ***p*** |
| **Ventricles – Total** | 21'185 (16'714, 78.9%) | 19'612 (16'045, 81.8%) | 22'560 (17'682, 78.4%) | *0.638* | **1.93** (1.52, 78.7%) | 1.86 (1.33, 71.5%) | 1.99 (1.71, 85.7%) | *0.813* | - | - | - | - |
| **Lateral ventricle – Total** | 18'430 (15'999, 86.8%) | 17'214 (15'961, 92.7%) | 19'493 (16'476, 84.5%) | *0.704* | **1.67** (1.45, 86.4%) | 1.62 (1.32, 81.7%) | 1.72 (1.59, 92.2%) | *0.853* | **83.20** (6.84, 8.2%) | 82.90 (8.22, 9.9%) | 83.46 (5.62, 6.7%) | *0.826* |
| *Lateral ventricle – Frontal horn* | 6'124 (5'139, 83.9%) | 6'002 (5'667, 94.4%) | 6'231 (4'816, 77.3%) | *0.905* | **0.55** (0.45, 80.4%) | 0.56 (0.47, 83.0%) | 0.55 (0.44, 80.6%) | *0.908* | **28.10** (4.70, 16.7%) | 28.75 (4.33, 15.0%) | 27.54 (5.07, 18.4%) | *0.491* |
| *Lateral ventricle – Body* | 4'852 (4'798, 98.9%) | 4'805 (5'435, 113.1%) | 4'894 (4'348, 88.8%) | *0.960* | **0.44** (0.43, 97.0%) | 0.45 (0.45, 99.3%) | 0.43 (0.43, 98.1%) | *0.925* | **21.12** (4.93, 23.3%) | 21.54 (6.02, 28.0%) | 20.75 (3.91, 18.8%) | *0.671* |
| *Lateral ventricle – Atrium* | 5'499 (5'464, 99.4%) | 4'547 (3'756, 82.6%) | 6'333 (6'627, 104.6%) | *0.381* | **0.50** (0.51, 102.6%) | 0.43 (0.31, 73.2%) | 0.56 (0.64, 114.6%) | *0.490* | **24.20** (5.96, 24.6%) | 22.99 (4.66, 20.3%) | 25.25 (6.88, 27.2%) | *0.309* |
| *Lateral ventricle – Occipital horn* | 1'094 (1'014, 92.7%) | 1'175 (1'257, 106.9%) | 1'022 (779, 76.2%) | *0.688* | **0.099** (0.092, 92.6%) | 0.11 (0.11, 103.0%) | 0.089 (0.069, 77.3%) | *0.512* | **4.95** (2.61, 52.6%) | 5.35 (2.76, 51.7%) | 4.61 (2.50, 54.3%) | *0.449* |
| *Lateral ventricle – Temporal horn* | 860 (619, 72.0%) | 685 (401, 58.4%) | 1'013 (741, 73.1%) | *0.151* | **0.080** (0.061, 76.5%) | 0.066 (0.038, 56.6%) | 0.091 (0.075, 82.3%) | *0.271* | **4.83** (2.74, 56.7%) | 4.28 (1.95, 45.7%) | 5.31 (3.27, 61.5%) | *0.309* |
| **Third ventricle** | 1'134 (808, 71.2%) | 847 (372, 43.9%) | 1'386 (998, 72.1%) | *0.068* | **0.10** (0.076, 73.0%) | 0.083 (0.036, 43.3%) | 0.12 (0.096, 78.5%) | *0.158* | **6.02** (1.79, 29.7%) | 5.42 (1.94, 35.8%) | 6.54 (1.51, 23.1%) | *0.088* |
| **Fourth ventricle – Total** | 1'620 (345, 21.3%) | 1'551 (295, 19.0%) | 1'681 (383, 22.8%) | *0.309* | **0.15** (0.033, 22.2%) | 0.15 (0.029, 19.0%) | 0.15 (0.037, 25.4%) | *0.684* | **10.78** (5.59, 51.9%) | 11.67 (6.56, 56.2%) | 10.00 (4.67, 46.7%) | *0.422* |
| *Fourth ventricle – Apex* | 163 (45.6, 28.0%) | 153 (44.1, 28.8%) | 171 (46.5, 27.1%) | *0.277* | **0.015** (0.0042, 28.4%) | 0.015 (0.0043, 28.7%) | 0.015 (0.0043, 29.0%) | *0.975* | **1.11** (0.67, 60.7%) | 1.19 (0.78, 66.0%) | 1.04 (0.58, 55.6%) | *0.570* |
| *Fourth ventricle – Lateral recess* | 199 (46.3, 23.3%) | 195 (45.7, 23.4%) | 201 (48.0, 23.8%) | *0.715* | **0.018** (0.0041, 22.6%) | 0.019 (0.0041, 21.4%) | 0.018 (0.0042, 23.8%) | *0.328* | **1.31** (0.66, 50.2%) | 1.44 (0.76, 53.1%) | 1.19 (0.54, 45.7%) | *0.311* |
| *Fourth ventricle – Obex* | 174 (109, 62.7%) | 180 (109, 60.7%) | 170 (113, 66.6%) | *0.813* | **0.016** (0.0097, 60.9%) | 0.017 (0.010, 58.5%) | 0.015 (0.0094, 64.2%) | *0.429* | **1.14** (0.85, 74.2%) | 1.33 (0.98, 73.8%) | 0.97 (0.69, 71.1%) | *0.244* |

| | | | | | | | | | | | | |
|---|---|---|---|---|---|---|---|---|---|---|---|---|
| *Fourth ventricle – Fastigium* | 199 (53.9, 27.0%) | 194 (54.5, 28.1%) | 204 (54.7, 26.8%) | *0.635* | **0.018** (0.0048, 26.4%) | 0.019 (0.0050, 26.5%) | 0.018 (0.0047, 26.8%) | *0.477* | **1.31** (0.66, 50.4%) | 1.45 (0.77, 53.5%) | 1.19 (0.54, 45.5%) | *0.303* |
